# Using Genomic Context Informed Genotype Data and Within-model Ancestry Adjustment to Classify Type 2 Diabetes

**DOI:** 10.1101/2024.09.12.24313579

**Authors:** Eric J Barnett, Yanli Zhang-James, Jonathan Hess, Stephen J. Glatt, Stephen V Faraone

## Abstract

Despite high heritability estimates, complex genetic disorders have proven difficult to predict with genetic data. Genomic research has documented polygenic inheritance, cross-disorder genetic correlations, and enrichment of risk by functional genomic annotation, but the vast potential of that combined knowledge has not yet been leveraged to build optimal risk models. Additional methods are likely required to progress genetic risk models of complex genetic disorders towards clinical utility. We developed a framework that uses annotations providing genomic context alongside genotype data as input to convolutional neural networks to predict disorder risk. We validated models in a matched-pairs type 2 diabetes dataset. A neural network using genotype data (AUC: 0.66) and a convolutional neural network using context-informed genotype data (AUC: 0.65) both significantly outperformed polygenic risk score approaches in classifying type-2 diabetes. Adversarial ancestry tasks eliminated the predictability of ancestry without changing model performance.

## Introduction

Over the past decade, many studies have developed regression-based and machine learning models for predicting the risk of complex genetic disorders^1^. Despite progress, their performance still falls short of the estimated limits based on heritability ^2^. Meanwhile, efforts to identify functional genome components and environmental factors influencing heritable disorders have advanced. In polygenic disorders, where heritability is spread across many genes, leveraging current genomic knowledge could improve risk estimation. One study showed that using trait-specific functional priors increased polygenic prediction accuracy by 5% ^3^. However, machine learning models have yet to fully utilize genomic knowledge, requiring new methods to handle the changes in data dimensionality and characteristics.

As an example, we focused on type 2 diabetes mellitus (T2D), which afflicts 6.3% of the world’s population ^4,5^. Genome-wide association studies (GWASs) of T2D have been successful in identifying hundreds of genetic variants ^6,7^. Studies using machine learning models to classify people with and without T2D have been less successful; model performances are lower than expected relative to the heritability of T2D and the classification performance of diseases with similar heritability ^8-10^. This poor performance may stem from several factors. One possibility is that the genetic risk for T2D is more complex, involving numerous small-effect loci across the genome, unlike type-1 diabetes, where risk is concentrated in a single region ^11^. If so, more sophisticated models may be needed to capture this complexity.

The complexity of machine learning models has its drawbacks ^12^. A complex model can capture patterns specific to the training data. If these patterns are more effective at classification than the real effects, the model may prioritize them, leading to “overfitting.” This means the model performs well on training data but struggles with unseen data that lack the same patterns. One example of a model learning patterns specific to training data and failing to generalize is when it is trained with population stratification ^13^. In population stratification, cases and controls in the training data have systematic ancestry differences. A model may use these variant frequencies between ancestry groups to classify cases and controls, but it ends up classifying ancestry rather than disease. When tested on data without the same ancestry imbalance, the model’s disease classification performance will drop.

Although ancestry can confound analyses, modeling ancestry alongside genotype data can improve prediction ^14,15^due to differing sampled case/control ratios within different ancestries or the interaction between genetic risk and a factor in which ancestry acts as a proxy. While these factors are undoubtedly important for clinical implementations of genetic risk models, the goal of many studies, including ours, is to better understand the genetic basis of the disorder, which motivates efforts to minimize the confounding effects of ancestry on genetic risk modeling.

In GWAS and polygenic risk score models, population stratification is typically controlled by including principal components as covariates to assess associations and derive risk scores ^16^. Most ancestry information is thought to be captured in the top principal components, so removing their effects allows the model to detect true disease risk. However, linear adjustments may not fully address ancestry-specific differences in allele frequencies. Since many machine learning models can learn non-linear relationships, it’s crucial to account for potential non-linear ancestry effects. In this work, we tackle this by using a novel within-model adjustment that reverses the gradient during gradient descent ^17^. This “adversarial task” helps the model unlearn ancestry, reducing its reliance on ancestry confounds.

One cause of overfitting complex models is the high number of parameters in a model ^18^. Many machine learning models can achieve near-perfect accuracy on randomly labeled data if the number of parameters far exceeds the number of samples ^19^. This is particularly relevant in GWAS studies, where genetic variants can number in the millions, while study participants are far fewer. A common way to reduce overfitting is to limit the number of variants used ^18^, but this may hinder model performance if true risk variants are excluded.

In this study, we developed a novel classification model using convolutional neural networks (CNNs) to limit the number of model parameters without removing genetic variants. CNNs are widely used in image recognition due to their ability to learn local patterns and efficiently reduce high-dimensional features. Several studies have shown the effectiveness of CNNs on genetic data, focusing on disease classification or annotation prediction based on genotype values ^20,21^. Here we take a novel approach by creating a CNN guided by genetic annotations, aiming to produce a low-dimensional representation of disease-relevant genetic variants. We hypothesized that training a CNN informed by genomic context would improve classification performance relative to current approaches. We also aimed to test whether using gradient reversal layers to create adversarial multi-task networks could control for ancestry confounding while preserving disorder classification accuracy.

## Methods

### Data Acquisition and Preprocessing

We obtained genotype and phenotype data from the UK Biobank ^22^, a prospective cohort study of over 500,000 individuals in the United Kingdom. The genotype data were generated by a combination of the UK Biobank Axiom array and UK BiLEVE Axiom array. 1,037 sample outliers, multi-allelic single nucleotide polymorphisms (SNPs), and SNPs with a minor allele frequency (MAF) < 1% were removed, resulting in 641,018 SNPs ^23^. These were used to impute ungenotyped SNPs, leading to a dataset of 73,355,667 variants. For our analyses, based on our training subset, we removed all SNPs with MAF < 1%, all SNPs with > 5% missingness, all individuals with > 5% missingness, and one member of any estimated kinship equal to or closer than second-degree relatives using Plink. We randomly split the data into training (70%), validation (15%), and testing subsets (15%). In each subset, we performed 1:1 case-control matched pairing based on age and sex using *MatchIt* ^24,25^. The training, validation and test subsets had 55,168, 11,712, and 11,648 records, respectively.

### Context Informed Data Matrix (CID)

#### Model Overview and Input

Simplified diagrams of model architectures, inputs, and outputs are shown in Figure 1. An overview of the models used in our analysis is shown in Table 1. The genotype input used in our NN models is the number of minor alleles at each SNP for each person, ranging from 0, when neither chromosome contained the alternate allele, to 2, when both chromosomes contained the alternate allele. The additional input used in our CNN models is a CID, which contains each SNP along with information about that SNP. The CID’s columns represent each SNP; rows contain the genomic annotations and disorder risk values for each SNP. Figure 2 shows a simplified CID.

**Table 1.** Overview of Machine Leaming Models.

| Model Name | Architecture Diagram | Model Input | Model Type | Normal Task(s) | Modified Task(s) |
| --- | --- | --- | --- | --- | --- |
| PC NN | Figure 1a | PCs | NN | T2D | --- |
| Geno-PC | Figure 1b | Genotype | NN | PCs | T2D from PCs (stop gradient) |
| Geno-PC-T2D | Figure 1c | Genotype | NN | T2D, PCs, T2D from PCs | --- |
| Geno-T2D Track-PC | Figure 1d | Genotype | NN | T2D | PCs (stop gradient) |
| Geno-T2D Adv-PC | Figure 1e | Genotype | NN | T2D | PCs (adversarial) |
| CID-T2D Track-PC | Figure 1f | CID and genotype | CNN | T2D, T2D from genotype | PCs (stop gradient) |
| CID-T2D Track-PC/Geno | Figure 1g | CID and genotype | CNN | T2D from CID | T2D from genotype (stop gradient), PCs (stop gradient) |
| CID Specific | Figure 1h | CID and genotype | CNN | T2D from CID | T2D from genotype (adversarial), PCs (adversarial) |
| Geno Specific | Figure 1i | CID and genotype | CNN | T2D from genotype | T2D from CID (adversarial), PCs (adversarial) |
| Combined Specific | --- | Output from final intermediate layers in Specific models | NN | T2D | --- |

**Figure 1.**
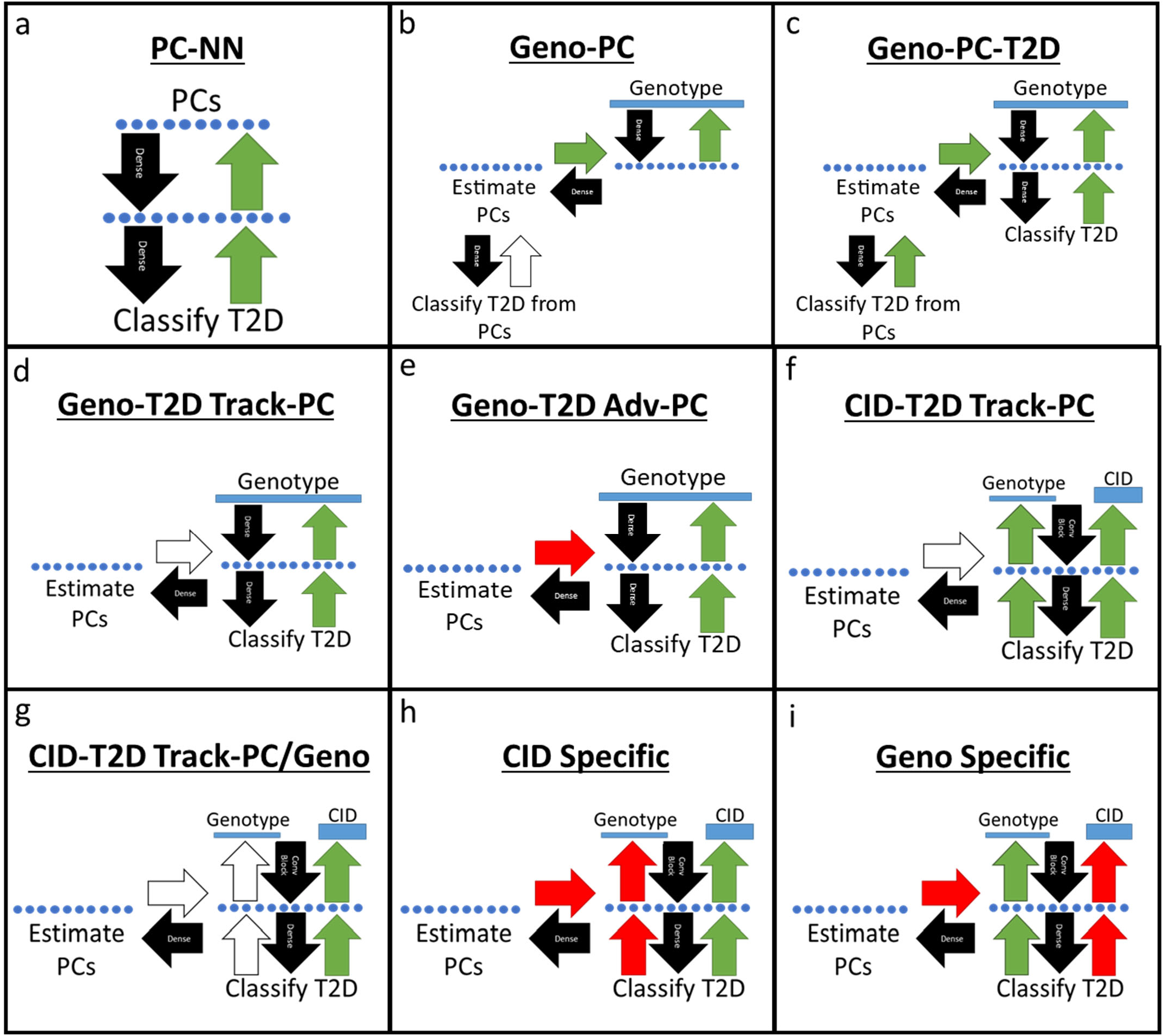
Simplified model architecture diagrams. The Black arrows represent the layers that connect each input to each output. The green arrows represent the positive feedback from backpropagation that aims to minimize error. The unfilled/white arrows represent stop gradient layers, which prevent the task from changing the weights in all layers upstream from the stop gradient layer. Red arrows represent gradient reversal layers of adversarial tasks, which reverse the direction of the weight changes and maximize loss for the task in any layer upstream from the gradient reversal layer. Note that some models, like panels f vs g and h vs I, differ only in the way gradients flow during backpropagation, illustrated by the color of the arrows. The blue rectangles represent the input data and the blue circles represent the transformed input that has been reduced to fewer dimensions. Task specific output layers (not shown) are present for each task in each model.

**Figure 2.**
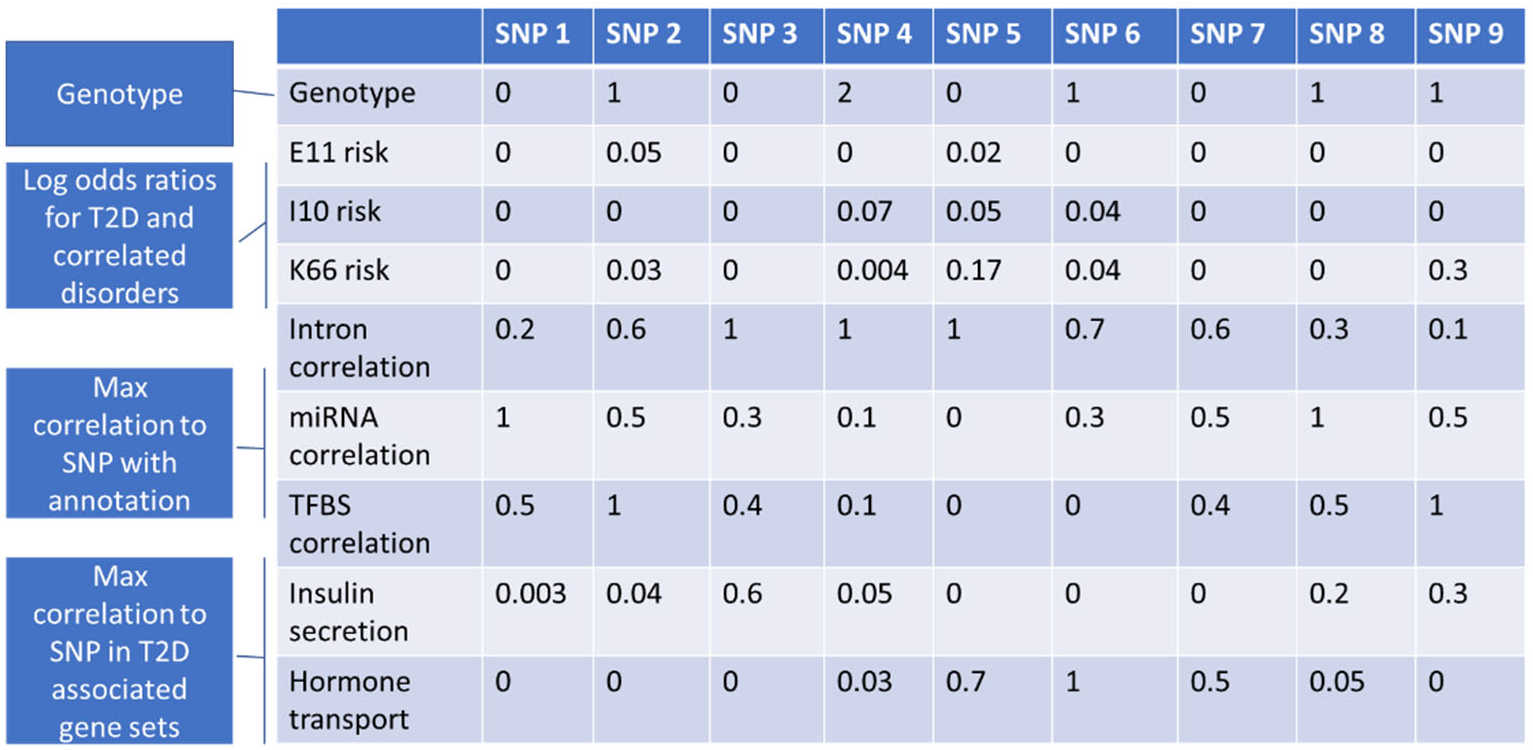
An illustrative example of a portion of a context informed data matrix (CID). A CID is constructed for each person within the study. For each person, the annotation values are multiplied by the allele count at each SNP and the resulting individualized annotation matrix is used as input into machine learning models.

For each individual, the CID, which largely contains information about each SNP that does not change between individuals, is multiplied by the genotype values of the individual prior to model training. For all models, we used a binary variable representing the presence of the International Statistics Classification of Disease 10^th^ revision (ICD-10) code for T2D, E11, for each individual as the label to be predicted. Our models output values from 0 to 1 with the goal of minimizing the error in predicting the label for each individual. The hyperparameter optimization ranges and values are in Table 3.

#### Genomic Annotations

The genomic annotations provided information on whether the SNP occurs at a location known to be an miRNA binding site, DNase hypersensitivity site, CPG island, gene, intron, 5’ untranslated region (UTR), 3’ UTR, splice site, promoter, or transcription factor binding site using the *AnnotationHub* ^26^ and *VariantAnnotation* ^27^ packages in R. We did this by loading the SNP locations and annotation ranges, and then finding the overlaps between the two. Any SNP that had an overlapping location with an annotation was coded as a 1 for that annotation or was otherwise coded as a zero. We included whether the SNP was a coding variant using the same overlap method.

We also added annotations indicating whether the SNP was within the range of each of the 20 gene sets most associated with T2D among gene sets in the lowest 10% standard error in *MAGMA* gene set analysis ^28^. We used the lowest 10% standard error threshold to ensure that the resulting gene set annotations were dense due to using larger gene sets and gene set association was consistent in the training subset as determined by MAGMA gene set analysis. The results of *MAGMA* gene set analysis on the training subset can be found in Supplementary Table 4.

#### Risk Values

To give the model information about documented disease risk associations, we added the log odds ratios for each SNP from an external GWAS on T2D^6^ to the CID. Only SNPs present in both the UK Biobank data and the external GWAS were included in the model. To reduce the computational cost of this analysis, we further reduced the number of SNPs for ML models by only including SNPs that were associated with T2D in the external GWAS at p ≤ 0.01. This reduction left us with 11,730 SNPs for machine learning models. We also included odds ratios from correlated disorders based on genetic correlations calculated by GWAS Atlas.^29^ From the list of the 100 most correlated traits with T2D, we selected traits that had ICD-10 codes available in the UK Biobank data. The included traits were overweight and obesity, disorders of lipoprotein metabolism and other lipidemias, essential hypertension, chronic ischemic heart disease, cholelithiasis, angina pectoris, other disorders of the urinary system, and pain in the throat and chest. We performed GWASs on these traits using genotyped UK Biobank individuals that were not included in our machine learning models. The top 10 principal components (PCs) were used as covariates in the model to adjust for population stratification. We included the log odds ratios calculated from these GWASs in the CID.

#### Correlation to Annotation

It is likely that, for most SNPs, the risk associated with the SNP is due to another genetic change that is associated with the SNP through linkage disequilibrium^30^. To account for this, we added an annotation for each previously described binary annotation that indicates the maximum squared correlation each SNP has to a SNP with the binary annotation. Correlations were calculated using *Plink* ^31^. In cases where the SNP has the annotation, the correlation is 1 and the resulting annotation value is identical to the original annotation value.

#### Model Hyperparameter Optimization

Within *TensorFlow* ^32^, we used the *KerasTuner* ^33^ framework to optimize the hyperparameters of our models. Using the Hyperband search algorithm, we searched for the optimal number of genomic convolutional blocks, number of filters within each block, filter and pool width within each block, number of dense layers, number of nodes and dropout rate within each layer, gradient reversal weights, L2 regularization presence and factor, number of epochs, and learning rates. The objective of the search algorithm was to maximize the area under the receiver operating characteristic curve (AUC) in the validation subset for classification tasks and maximize the validation subset R^2^ for regression tasks.

#### Detecting and Adjusting Ancestry Confounding with Principal Components and Adversarial Learning

ML models use all information available to them to produce the best performance possible. This can result in models that perform well mainly due to a confounding variable. To address confounding by ancestry, we ran a principal components analysis using the SNPs that were not included in our ML models nor correlated with the SNPs (r^2^ < 0.2) that were used in the ML models. We extracted the top 10 PCs, as is frequently used for ancestry inference ^16^, and used them as labels in several models.

To establish whether population stratification was an issue, we built the “PC NN” model (Figure 1a) using the ancestral principal components to predict T2D status. To determine if the subset of SNPs used as predictors in our model could recreate the principal components within the model, we used the genotype data as input in a neural network model that predicted the PCs (“Geno-PC” model, Figure 1b). We determined the effectiveness of all model architectures in estimating the PCs with mean squared error (MSE) and R^2^. In models that had a positive R^2^ value, we also tracked T2D classification AUC from the PC estimates.

To test if a neural network could classify T2D and estimate PCs using the same layers, we designed a multi-task model with both tasks sharing all layers except output layers (“Geno-PC T2D” model, Figure 1c). An additional task classifying T2D from the PC estimates was used to compare classification performance between the PC estimates and the true PCs.

To test whether, in classifying T2D from genotype data, our model inadvertently uses PC-like ancestral information, we built a multi-task model where one task classified T2D from the genotype input and another task estimated PCs (“Geno-T2D Track-PC” model, Figure 1d) from the output of the dense layers. In this model, we used a stop gradient layer between the PC estimation task and the dense layers, which stops backpropagation. By using this layer, we stopped training such that the task estimating PCs did not influence the dense layers upstream from the layer unique to their tasks. This strategy allows us to track the performance of a task, in this case, PC estimation, without interfering with shared layer training. If the PC estimation task was predictive, this would suggest the model classifying T2D from genotype data used ancestral information that could be directly transformed into PC estimations.

To test whether we could teach the network not to use ancestry information when predicting T2D, we built a multi-task model (“Geno-T2D Adv-PC” model, Figure 1e) with the same two tasks as the previous model but replaced the stop gradient layer with a gradient reversal layer, a technique initially developed in the domain adaptation field of machine learning ^17^. This layer reverses the direction of gradients in gradient descent, thereby directing the weights in the layers upstream of the gradient reversal layer to adjust in a way that maximizes the error in the task, instead of the typical minimization. Unlike the fast gradient sign method, which creates “adversarial examples” by using gradients to create a new example that maximizes loss, the gradient reversal layer creates a continuous “adversarial task” by reversing the gradient from back propagation from the task specific layers of the selected task throughout model training. The “adversarial task” directs the model to find ancestry-invariant patterns by forcing the shared layers to learn features that make it hard to predict ancestry. This adversarial learning setup effectively makes the representations non-informative with respect to ancestry. Layers that are downstream of the gradient reversal layer still adjust weights in a way that minimizes error in the task. This means the adversarial task still attempts to minimize error and accurately estimate PCs in the layers that are unique to the task, thereby leaving adjustment of shared layers as the only option in maximizing error. We used this architecture to remove any PC-like ancestry information from the shared layers. If the model is unable to estimate PCs, it suggests the ancestry information present within the PCs is not being used in the shared layers.

#### Convolutional Neural Network Model Architecture for Genomic Data

In all our CNN models, we used the matrices as input into genomic convolutional blocks, which are repeating units within the model that contain a combination of 1D convolutional layers with ReLU activation functions, pooling layers, and batch normalization layers. Within convolutional layers, the convolutional filters require spatial invariance, meaning a signal on one part of the two-dimensional data structure means the same thing as the same signal anywhere else in the data structure. This is true for images, for which convolutional layers were originally developed. In contrast, the height dimension of the context-informed data matrix used as input in our model, which is the annotation information at each SNP location, has no spatial meaning, and filters at different heights could mean vastly different things, which violates spatial invariance. To address this issue, we set the height of each convolutional filter equal to the total number of rows present in the matrix to assure that output signals from one part of the matrix were equivalent to those from another part. Given that spatial invariance is violated along the genome, with the columns representing SNPs in the CID, we set the hyperparameter selection range of convolutional width to 1-20. This range was used to allow the model to set the convolutional width to 1 if having convolutional filters containing more than a single SNP were disruptive to model performance due to the violation of spatial invariance, while also allowing for wider convolutional filters if that was beneficial to model performance. After each 1D convolutional layer, the output was fed into a batch normalization layer. After the genomic convolutional blocks, the output was used as input into one or more dense layers with ReLU activation functions, depending on hyperparameter optimization. Finally, we used a dense layer with a sigmoid activation function to produce an output prediction of the T2D status for each person. We used the Adam optimizer and binary cross-entropy loss function to train our models.

To test whether genotype-only models and context-informed genotype models can share the same features, we trained a CNN model that had two input sources that shared intermediate layers but have separate input-specific T2D classification tasks (“CID-T2D Track-PC” model, Figure 1f). This means that the two input sources undergo the same transformations in the shared layers, but the output of those transformations is different because the input sources are different. The inputs into this model differ in that the CID input contains annotation information for each SNP whereas the genotype input has copies of the genotype data such that it is the same shape as the CID input to allow layer sharing. The input specific output layers then optimize T2D classification based on the result of the transformations in each input source. One input was the CID as previously described. The second input was the genotype data portion of the CID replicated such that it had the same dimensions as the CID. Both inputs shared all layers except the output layer specific to each input/task. If both tasks can classify T2D, it would suggest that there are similarities in the patterns used in the two input types. A PC estimation task with a stop gradient layer was used to track whether ancestry information was used within the model.

To test whether overlaps in the patterns found and used in both tasks are present when only training the CNN with CID input, we trained another CNN model (“CID-T2D Track-PC/Geno” model, Figure 1g). In this model, we used the same base structure as the “CID-T2D Track-PC” model but used a stop gradient layer prior to the output layer of the genotype-only model, thereby preventing the task from training layers besides the output layer unique to the task. If the task is still able to classify T2D diagnosis with these constraints, it suggests that at least some of the similarities in the patterns used for both tasks are used even when not directed to find shared patterns.

To test whether the CNN with CID input can find patterns that the genotype-only model cannot use to classify T2D, we built a third CNN model (“CID Specific” model, figure 1h). In this model, the stop gradient layers in the “CID-T2D Track-PC/Geno” model were replaced with gradient reversal layers. This effectively forces the shared layers away from any patterns that could be used by the genotype only input to classify T2D. Likewise, the shared layers are forced away from using ancestry information that could be used to estimate PCs. If the CID input can classify T2D using the same layers as the genotype only input, it would suggest that the CID input can model patterns that the genotype only input is not able to model. To test whether the CNN with genotype input can find patterns the model with CID input cannot, we built a similar model (“Geno Specific” model, Figure 1i) that switched the main and adversarial tasks of the “CID Specific” model.

To test whether the “Specific” models contained non-overlapping T2D risk information we built a model that used a combination of the risk features generated by the “Specific” models to predict T2D. To do this, we concatenated the output from the final intermediate layer in both models. This combined output was used as input into a neural network model (“Combined Specific” model) with the task of classifying T2D diagnosis. We compared the performance of this combined model to the performance of the “Specific” models individually.

#### Logistic Regression Models and Model Comparison

We fit logistic regressions using multiple PRS methods on all available SNPs comparisons. For standard PRS, we used Plink to prune correlated SNPs and calculate polygenic risk scores (PRSs)^31^. We also calculated PRS scores using LDpred2 and PRS-CS^34,35^. We adjusted all PRSs for ancestry by regressing the PCs on the PRSs and keeping the residual. We used this residual in the generalized linear model (glm) function within the base stats package in *R* to fit a logistic regression using only the training subset. We predicted the test subset T2D status and the performance within the test subset to compare to other models. We used AUC to measure performance in the test subset. AUC confidence intervals were computed with 2,000 stratified bootstrap replicates using the *pROC R* package ^36^. We also used the *pROC* package to compare the performance between models with DeLong’s test for two correlated ROC curves.

#### Feature Importance and Generalization Analyses

We used Integrated Gradients on our trained NN and CNN models to identify the features that were most influential in separating cases and controls.^37^ The output from Integrated Gradients is a value for each input feature for each person representing how much that feature contributed to the prediction of that person’s T2D status. To estimate overall feature importance, we compared the mean attributions in cases and controls for each feature using t-tests. We adjusted for multiple comparisons in our feature importance analyses using Bonferroni correction. There is significant and reasonable concern that the features used in ML algorithms are somewhat specific to the data used to train the model^18^. To test for feature importance generalizability, we calculated the correlations between the probit transformed feature importance p-values estimated from the training subset and testing subset. We compared these correlations between models to compare the feature importance generalizability. To compare these results to more traditional approaches, we also calculated correlations between the probit transformed GWAS p-values generated in the training subset and testing subset. We used a t-test to determine if the groups of SNPs with feature importance p-values less than 0.05 had significantly different GWAS association p-values compared to the non-significant group.

## Results

In the following, “genotype data” refers to the set of genotypes used as input to the models, “GWAS PCs” refers to ancestrally informative PCs estimated from the full set of GWAS SNPs and “ML PC estimates” refers to PCs estimated in machine learning models using genotype data as input. Table 2 shows the performance of all models on the test subset. Descriptions of each model’s results can be found in the supplementary results.

**Table 2.** Machine Leaming Model Results.

| Model Input | Model Type | Architecture Diagram | Task(s) | T2D Classification AUC (95% CI) | Estimated PC MSE | Estimated PC R<sup>2</sup> | T2D Classification from Estimated PCs AUC (95% CI) | T2D Classification from Alternate Input |
| --- | --- | --- | --- | --- | --- | --- | --- | --- |
| PCs | NN | Figure 1a | T2D | 0.56 (0.54 – 0.56) | --- | --- | --- | --- |
| Genotype | NN | Figure 1b | PCs, T2D from PCs (stop gradient) | --- | 0.27 | 0.73 | 0.55 (0.54 – 0.56) | --- |
| PC adjusted PRS | LR | --- | T2D | 0.57 (0.56 – 0.58) | --- | --- | --- | --- |
| PRS-CS | LR | --- | T2D | 0.59 (0.58 – 0.60) | --- | --- | --- | --- |
| LDpred2 | LR | --- | T2D | 0.63 (0.62 – 0.64) | --- | --- | --- | --- |
| Genotype | NN | Figure 1c | T2D, PCs, T2D from PCs | 0.66 (0.65 – 0.67) | 0.38 | 0.62 | 0.56 (0.55 – 0.57) | --- |
| Genotype | NN | Figure 1d | T2D, PCs (stop gradient) | 0.65 (0.64 – 0.66) | 1.70 | < 0 | --- | --- |
| Genotype | NN | Figure 1e | T2D, PCs (adversarial) | 0.66 (0.65 – 0.67) | 2.32 | < 0 | --- | --- |
| CID<br>Alternate: Genotype | CNN | Figure 1f | T2D, T2D from genotype, PCs (stop gradient) | 0.62 (0.61 – 0.63) | 0.69 | < 0 | --- | 0.63 (0.62 – 0.64) |
| CID<br>Alternate: Genotype | CNN | Figure 1g | T2D from CID, T2D from genotype (stop gradient), PCs (stop gradient) | 0.65 (0.64 – 0.66) | 0.66 | < 0 | --- | 0.54 (0.53 – 0.55) |
| CID<br>Alternate: Genotype | CNN | Figure 1h | T2D from CID, T2D from genotype (adversarial), PCs (adversarial) | 0.59 (0.58 – 0.60) | 3.30 | < 0 | --- | 0.50 (0.50 – 0.50) |
| Genotype<br>Alternate: CID | CNN | Figure 1i | T2D from genotype, T2D from CID (adversarial), PCs (adversarial) | 0.57 (0.56 – 0.58) | 3.37 | < 0 | --- | 0.50 (0.49 – 0.51) |
| h + i intermediate layer output | NN | --- | T2D | 0.61 (0.60 – 0.62) | --- | --- | --- | --- |

**Table 3.** Hyperparameter Optimization Ranges.

| Optimization Task | Optimization Range |
| --- | --- |
| Number of Convolutional Blocks | 1 - 3 |
| Number of Convolutional Filters | 10 - 100 |
| Convolutional Filter Width | 1 - 20 |
| Pooling Width | 1 - 20 |
| Number of Dense Layers | 1 - 3 |
| Nodes Within Dense Layer | 5 - 100 |
| L2 regularization factor | 0 - 1e-4 |
| Dropout Rate | 0 - 0.5 |
| Gradient Reversal Weight | 0.001 - 0.2 |
| Learning Rate | 1e-6 - 1e-3 |

### Adversarial Ancestry Adjustment

Two of our models (shown in Figures 1a and 1b) showed that both GWAS PCs and ML PC estimates were significantly predictive of T2D. The prediction of T2D using GWAS PCs and the prediction of T2D using ML PCs estimates were not significantly different (p = 0.65, Delong’s test for two correlated ROC curves). This illustrates the issue that within model correction may be necessary to avoid using potentially confounding ancestry information in classification modeling. Models that used adversarial tasks for ancestry (shown in Figures 1e, 1h, and 1i) all increased the ML PC estimates MSE as intended, ensuring that the model could not accurately recreate GWAS PCs and therefore could not use the ancestry information within those PCs to improve the classification of T2D. All models with an adversarial ancestry task had an R^2^ < 0 for the PC estimation task, indicating that the model’s predictions are worse predictions than using the mean.

### Classification Model Performance and Comparison

The best performing NN model was the “Geno-T2D Adv-PC” model (Figure 1e), with an AUC of 0.66 (95% CI: 0.65 – 0.67). The best performing CNN model was the “CID-T2D Track-PC/Geno” model (Figure 1g) with an AUC of 0.65 (95% CI: 0.64 – 0.66). In comparison, the best performing PRS method used LDpred2 to predict T2D with an AUC of 0.63 (95% CI: 0.62 – 0.64). The best performing CNN and NN models were both significantly more predictive than the best PRS method (p = 0.009 and p = 0.001, Delong’s test for two correlated ROC curves) despite using only 11,730 SNPs compared to 979,891 SNPs used in the PRS methods.

### Feature Importance and Generalization Analyses

SNPs with significantly different mean attributions between cases and controls in the test subset for the “Geno-T2D Adv-PC”, “CID-T2D Track-PC/Geno”, and “CID Specific” models can be found in Supplementary Tables 1-3. In all 3 models, the significant SNPs based on feature importance did not have significantly different GWAS association p-values compared to the SNPs that were not significant based on feature importance. All of the methods we investigated showed statistically significant generalizability, shown by the correlations between association/feature importance p-values in the training and testing subsets. The association p-values of a GWAS using the training subset and a GWAS using the testing subset had a correlation of 0.19 (95% CI: 0.18 – 0.21). The feature importance p-values in the training and testing subsets in the “Geno-T2D Adv-PC” model had a correlation of 0.70 (95% CI: 0.69 – 0.71). The “CID-T2D Track-PC/Geno” model had a correlation of 0.63 (95% CI: 0.62 – 0.64). The “CID Specific” model had a correlation of 0.66 (95% CI: 0.65 – 0.67). All 3 ML models had significantly higher p-value correlations compared to GWAS.

## Discussion

Our findings show that incorporating genetic annotations for common variations provides new insights not seen in previous genomic machine learning models. We are the first to use convolutional layers with genomic context, and our results suggest these methods can find unique, generalizable risk patterns. This study is also the first to apply gradient reversal layers in genomic machine learning, proving useful for adjusting ancestry and testing hypotheses. We found that relying solely on the top 10 principal components is insufficient for removing ancestry-related confounding. Therefore, within-model control methods are essential to prevent ancestry confounding in predictive models.

In a large dataset of over 70,000 subjects, the “PC NN” model (Figure 1a) significantly predicted T2D using the top 10 PCs from a PCA based on SNPs not included or in linkage disequilibrium with SNPs used to estimate our models. The “Geno-PC” model (Figure 1b) showed that the genotype information we use as predictors can accurately estimate PCs. Taken together, these two models show that machine learning models can recreate ancestry information and use that information to classify T2D diagnosis and, likely, other disorders. The “Geno-PC-T2D” model (Figure 1c) tested this ancestry inference to disorder diagnosis pathway directly and resulted in T2D classification performance not significantly different from classifying directly from GWAS PCs. The “Geno-T2D Adv-PC” (Figure 1e) did not perform any worse than the “Geno-PC-T2D” model (Figure 1c), despite eliminating the latter model’s ability to estimate PCs. Since the layers within the Geno-T2D Adv-PC model were forced away from using ancestry information, it is likely that nodes of the model that had been occupied with ancestry inference were used to represent additional real risk features of T2D.

In traditional machine learning approaches, it’s difficult to know if ancestry influences a model. Our solution uses a subtask that estimates ancestry-adjusted principal components (PCs) from the output of layers used in the main classification task. This subtask cannot affect the shared layers’ weights due to a stop gradient layer. Without it, backpropagation would alter the main classification layers to improve PC estimation, encouraging the use of ancestry information. Instead, the stop gradient layer monitors whether ancestry data is used without altering the main network. If significant PC estimation occurs with the stop gradient, it indicates ancestry information is present in the shared layers. In this case, the stop gradient can be replaced by a gradient reversal layer, which flips the direction of weight changes from gradient descent in the upstream layers. This causes the model to maximize errors in PC estimation, avoiding ancestry use. This effect is shown by comparing the MSE in Figures 1d and 1e, where replacing the stop gradient with a gradient reversal layer increases the PC estimation MSE from 1.70 to 2.32. The strength of this adversarial task can be adjusted to eliminate ancestry information. If more detailed ancestry data is available, it can also be used similarly. Studies have shown that genetic risk models, primarily developed on people of European ancestry, do not generalize well to other ancestral groups ^38,39^. Adversarial ancestry tasks would reduce this discrepancy by finding ancestry-invariant patterns.

In the “CID-T2D Track-PC” model (Figure 1f), we trained a model to predict T2D with two types of inputs. The inputs were a “CID Input” that uses genomic context information and a “genotype input” that did not use genomic context information. The input types trained and used the same shared layers, apart from output layers that were not shared. The classification performance resulting from the two input types were similar. This suggests that the input types can find and use the same patterns within the data to classify T2D. This result was expected since the two input types share the same genotype information.

However, the CID-T2D Track-PC/Geno model (Figure 1g), which only allowed the CID input to train the shared layers, found that the CID input was significantly better at classifying T2D compared to genotype input that did not train the shared layers (p = 2e-16, Delong’s test for two correlated ROC curves). This shows that while the two input types share similarities in their ability to represent the risk of T2D, CID input can create unique risk representations that improve prediction beyond what can be represented by genotype data alone.

Our use of gradient reversal layers to create adversarial tasks in the “CID Specific” model and “Geno Specific” model further separated out input-dependent features. While both models’ performance declined relative to other CNN models, the inputs for the non-adversarial tasks still had AUCs that were statistically significant. In comparison, the adversarial task input (genotype input for the “CID Specific” model and CID input for the “Geno Specific” model) was unable to significantly classify T2D using the same shared layers used by the main input. This suggests that the risk representations within the models leading to T2D classification were specific to the non-adversarial input. Our feature importance analysis also suggests that the CNN models are finding different risk patterns compared to our NN models. There was only a 2% overlap in significantly predictive SNPs between the “Geno-T2D Adv-PC” and the “CID Specific” models.

One explanation of the unique risk representations is that they represent the same underlying risk features but are constructed in a way that can only be used with one input type. To test this theory, we trained the “Combined Specific” model, which combines the features of the final intermediate layers of the “CID Specific” and “Geno Specific” models. If the features from the “Specific” models were overlapping, one would expect the model using the features from both models to have the same classification performance. We found that the “Combined Specific” model was significantly better at classifying T2D compared to both individual models. This suggests that the risk representations found in the “Specific” models are at least in part specific to those input types. This suggests that some of the genetic risk for T2D can be transformed into higher-order risk features using genomic context (features from the “CID Specific” model) while other risk cannot be transformed with the set of genomic context information used in our analyses. Further investigation of the genetic variants and higher-order risk features used and developed in these models could reveal insights for future genetic risk modelling efforts.

The performances of our NN and CNN models improves on prior results. One study reported an AUC of 0.60 (95% CI: 0.59 – 0.61) when using a gradient boosted and LD-adjusted heuristic polygenic score and an AUC of 0.61 when using LDpred polygenic scoring ^40,41^. The same study reported an AUC of 0.58 in an LD-unadjusted polygenic risk score and an AUC of 0.58 when using traditional pruning and thresholding polygenic scoring methods, which is like our polygenic risk score methods and results. Another study that investigated many different models and feature encodings reported a maximum AUC in T2D across all models/encodings of 0.59^42^. We used the LDpred2 polygenic scoring method and saw an improvement compared to these prior results, with an AUC of 0.63 (95% CI: 0.62 – 0.64)^35^. Our best genotype (“Geno-T2D Adv-PC” model, AUC: 0.66, 95% CI: 0.65 – 0.67) and CID (“CID-T2D Track-PC/Geno” model, AUC: 0.65, 95% CI: 0.64 – 0.66) models are both improvements over these prior studies and our own polygenic scoring results. While these are significant improvements, they are small; further advances will be necessary to reach the goal of clinical utility.

Classification models may be useful for identifying genetic variants that predict T2D. Our feature importance analyses of the “Geno-T2D Adv-PC”, “CID-T2D Track-PC/Geno”, and “CID Specific” models had 352, 57, and 62 SNPs with statistically significant differences in mean attributions between cases and controls in the test subset (see Supplementary Tables 1-3). These SNPs did not have significant differences in association with T2D compared to the other SNPs used in the models. This suggests that information beyond the differing allele rates between cases and controls are driving the importance of the significant SNPs, which may be worthy of further investigation. They may also be specific to the models and data used in our study due to overfitting. We addressed generalizability of our models’ prediction in this study by presenting classification model performance only in a withheld testing subset of data that were not used to train or optimize the model. As an additional test of generalizability, we calculated the correlation of feature importance values calculated from the training subset to those calculated from the testing subset. Higher correlations in this analysis reflect higher generalizability of features to other data, further validating the reported significant SNPs in models with high correlations. All 3 of the models we tested showed higher correlations compared to those of a GWAS on the training and test subsets.

Several limitations may have reduced our models’ ability to classify T2D. Due to computational issues, we only used a small subset of the genetic variants. Using more variants might increase the CNNs’ ability to detect local patterns since the genetic variants would be spaced closer together and the distance between variants would be more uniform. Insufficient optimization of hyperparameters may have also limited the performance of our models. In more complex models, the hyperparameter space becomes too large to efficiently explore all possible solutions. Therefore, it is likely that our model architectures are not the optimal solution. Further exploration of the hyperparameter space could improve results.

The annotations used to create the CID may not be the best combination of information. Adjustments to the genomic context information might improve our results. There has been some evidence that population structure may not be fully captured by just the top PCs, so residual ancestry information could still influence the model ^43^. Our models’ ability to account for ancestry depends on the PCs used in the adversarial task. It is possible that other ancestry data are still included in the model and used in classification. Including more PCs as labels in adversarial tasks may further reduce the models’ reliance on ancestry information, minimizing the potential confounding effect of population stratification. On the other hand, our focus on minimizing ancestry information in the models may come at the cost of some predictive power. It is possible that some of the genetic risk features learned by the models in our study may be strengthened or weakened by the presence of gene-gene or gene-environment interactions present specifically in certain ancestry groups. Further study of ancestry-variant genetic risk will further our understanding and modeling of disorders like T2D. In addition, while our withholding of a test subset and large data set size allowed us to test within-distribution generalization of our models, the performance of our models trained on the UK Biobank data in other data sets is unknown. The most generalizable modeling strategy would likely be to train a single model using multiple data sets. Ideally, we would have tested the model from an entirely different data collection, but we do not have access to such data.

In summary, we have described novel CNN/NN architectures that combine genomic context-informed genotype data, and within model ancestry detection/adjustment. Our results indicate that this may be a useful direction for improving our ability to classify complex genetic disorders and detect SNPs that are significantly predictive of disease. While classification performance remains too low for clinical utility and earlier detection of T2D risk, incremental improvements such as those reported here may get to the point of clinical utility in the future.

## Supporting information

Supplementary Results

## Data Availability

This research was conducted using genotype and phenotype data from the UK Biobank resource under application number 60050. All other data are available from the authors upon request.

## Author Contribution Statement

E.J.B. and S.V.F. conceived and designed the models and analyses. E.J.B. implemented the models and performed the analyses with contributions from S.V.F., Y.Z.J., J.H., and S.J.G. E.J.B. wrote the paper. All authors discussed model design, results, and implications. All authors commented and edited the manuscript.

