## Supplementary Results for "Using Genomic Context Informed Genotype Data and Within-model Ancestry Adjustment to Classify Type 2 Diabetes"

The “PC NN” model (Figure 1a) had a small but significant level of predictive accuracy, with an AUC of 0.55 (95% CI: 0.54 – 0.56). The “Geno-PC” model, which used genotype input to estimate PCs (Figure 1b), which were calculated in a GWAS that did not include the genotype data used in the NN model, had an R^2^ of 0.73. The additional task that predicted T2D diagnosis from ML PC estimates in this model had a significant AUC of 0.55 (95% CI: 0.54 – 0.56). The PRS and PC adjusted PRS logistic regressions had AUCs of 0.60 (95% CI: 0.59 – 0.61) and 0.57 (95% CI: 0.56 – 0.58), respectively. These two models were significantly different (p = 2e-13).

The “Geno-PC T2D” Model (Figure 1c) was significantly predictive in all tasks. The T2D classification task had an AUC of 0.66 (95% CI: 0.65 – 0.67, the estimation of PCs task had an R^2^ of 0.62 and MSE of 0.38, and the T2D from PC estimates task had an AUC of 0.56 (95% CI: 0.55 – 0.57). The prediction of T2D from GWAS PCs and ML PC estimates based on genotype input was not significantly different (p = 0.35). The “Geno-T2D Track-PC” model (Figure 1d) had an AUC of 0.65 (95% CI: 0.64 – 0.66) for T2D classification and an R^2^ < 0, indicating that the model’s predictions on the PC estimates task are worse predictions than using the mean, and MSE of 1.70 for the PC estimation task. The “Geno-T2D Adv-PC” model (Figure 1e) had an AUC of 0.66 (95% CI: 0.65 – 0.67) for the T2D classification task, while the estimation of PCs task had an R^2^ < 0 and MSE of 2.32.

The “CID-T2D Track-PC” model (Figure 1f) had an AUC of 0.62 (95% CI: 0.61 – 0.63) for the T2D from CID input task and 0.63 (95% CI: 0.62 – 0.64) for the T2D from genotype only input task. The PCs estimation task with stop gradient layer designed to track but not influence PC usage in the shared layers had a R^2^ < 0 and MSE of 0.69.

The “CID-T2D Track-PC/Geno” model (Figure 1g) had an AUC of 0.65 (95% CI: 0.64 – 0.66) for the classification of T2D from CID input task. The AUC for classifying T2D from genotype only was 0.54 (95% CI: 0.53 – 0.55), which was significantly lower than the T2D from CID input task (p = 2e-16).

The “CID Specific” model (Figure 1h) had an AUC of 0.59 (95% CI: 0.58 – 0.60) for the T2D from CID input task. The classification of T2D from genotype-only input task was not significantly predictive (AUC = 0.50). The adversarial PCs estimation task had a R^2^ < 0 and MSE of 3.30. The “Geno Specific” model (Figure 1i) had an AUC of 0.57 (95% CI: 0.56 – 0.58) in its main task of classifying T2D from genotype input. This was a small but significant decrease in AUC compared to the “CID Specific” model (Figure 1h) (p = 0.0002). The “Combined Specific” model had an AUC of 0.61 (95% CI: 0.60 – 0.62). This was a significant increase compared to both the “CID Specific” (p = 8.4e-4) and “Geno Specific” (p = 1.8e-11) models.

**Supplementary Table 1: Geno-T2D Adv-PC Model Feature Importance T-test Results**

| **SNP** | **t_test_p_value** | **t_test_statistic** | **t_test_std_error** | **adjusted_p_value** |
| --- | --- | --- | --- | --- |
| rs2190095 | 1.35E-09 | 21.20475 | 2.44E-05 | 1.59E-05 |
| rs2157374 | 2.32E-09 | 21.49843 | 3.43E-06 | 2.72E-05 |
| rs4237133 | 3.75E-09 | 19.46848 | 1.12E-05 | 4.40E-05 |
| rs12887490 | 6.47E-09 | 23.80538 | 2.34E-05 | 7.58E-05 |
| rs12939056 | 9.60E-09 | 17.43501 | 7.14E-05 | 0.000113 |
| rs6941355 | 9.83E-09 | 17.59312 | 4.13E-05 | 0.000115 |
| rs9608698 | 1.28E-08 | 18.02013 | 1.11E-05 | 0.00015 |
| rs6060003 | 1.51E-08 | -19.8107 | 1.60E-05 | 0.000177 |
| rs4959424 | 1.87E-08 | 16.12801 | 1.88E-05 | 0.000219 |
| rs7841229 | 2.09E-08 | 16.3435 | 5.38E-05 | 0.000245 |
| rs1265158 | 2.13E-08 | -18.6625 | 9.91E-06 | 0.00025 |
| rs3094110 | 2.23E-08 | -15.8456 | 1.78E-05 | 0.000261 |
| rs4753426 | 2.40E-08 | 16.51468 | 1.24E-05 | 0.000281 |
| rs34802028 | 2.79E-08 | -15.3707 | 6.81E-06 | 0.000327 |
| rs6925389 | 2.93E-08 | -15.6343 | 3.85E-05 | 0.000343 |
| rs8103434 | 4.46E-08 | -15.7734 | 3.19E-05 | 0.000523 |
| rs849336 | 6.04E-08 | 14.98271 | 7.87E-05 | 0.000709 |
| rs3753474 | 7.28E-08 | -13.9743 | 3.61E-05 | 0.000854 |
| rs1653609 | 8.36E-08 | 17.26816 | 1.90E-05 | 0.00098 |
| rs11719375 | 8.81E-08 | 13.61865 | 2.42E-05 | 0.001034 |
| rs7451690 | 1.17E-07 | -13.2396 | 1.31E-05 | 0.001378 |
| rs7134021 | 1.21E-07 | -13.1981 | 6.14E-05 | 0.001417 |
| rs12565995 | 1.24E-07 | -13.4066 | 3.86E-06 | 0.00146 |
| rs7741306 | 1.39E-07 | 13.093 | 3.90E-05 | 0.001634 |
| rs2583930 | 1.39E-07 | 13.27353 | 0.0002 | 0.001636 |
| rs6686 | 1.41E-07 | -13.7607 | 3.06E-05 | 0.001658 |
| rs10881930 | 1.42E-07 | -13.1028 | 4.82E-05 | 0.001667 |
| rs10881996 | 1.44E-07 | 13.00855 | 9.54E-07 | 0.001684 |
| rs2071473 | 1.55E-07 | 16.95479 | 2.49E-05 | 0.001823 |
| rs2230657 | 1.61E-07 | -12.7865 | 3.70E-05 | 0.001887 |
| rs6486331 | 1.61E-07 | -14.5294 | 7.22E-06 | 0.001892 |
| rs34650965 | 1.66E-07 | 13.08554 | 7.82E-08 | 0.001951 |
| rs2956086 | 1.68E-07 | -12.8518 | 1.15E-05 | 0.001969 |
| rs58642235 | 2.10E-07 | 12.52789 | 1.78E-05 | 0.002459 |
| rs2293242 | 2.11E-07 | -13.3755 | 5.89E-05 | 0.002473 |
| rs8102445 | 2.13E-07 | -13.2878 | 2.84E-05 | 0.002495 |
| rs6453446 | 2.28E-07 | 12.40398 | 4.23E-05 | 0.002672 |
| rs10824271 | 2.30E-07 | 13.33682 | 5.63E-05 | 0.002693 |
| rs6933716 | 2.30E-07 | -12.443 | 3.79E-05 | 0.002701 |
| rs2440369 | 2.31E-07 | 13.50558 | 1.59E-05 | 0.00271 |
| rs3791710 | 2.32E-07 | -12.3029 | 3.51E-06 | 0.002716 |
| rs3846662 | 2.32E-07 | -16.1139 | 5.26E-05 | 0.002727 |
| rs2488597 | 2.36E-07 | -12.8547 | 8.31E-06 | 0.00277 |
| rs3783627 | 2.43E-07 | 12.92388 | 1.41E-05 | 0.002854 |
| rs7034165 | 2.67E-07 | 13.60427 | 1.88E-06 | 0.003128 |
| rs6496746 | 2.67E-07 | 12.96993 | 6.90E-05 | 0.003137 |
| rs10840038 | 2.72E-07 | -12.1497 | 2.77E-05 | 0.003189 |
| rs2071339 | 2.75E-07 | 12.14039 | 3.10E-05 | 0.003223 |
| rs9874893 | 2.81E-07 | 12.79205 | 3.12E-05 | 0.003294 |
| rs238239 | 2.85E-07 | 12.12092 | 4.10E-05 | 0.003338 |
| rs12916535 | 2.88E-07 | -12.0198 | 1.29E-05 | 0.003374 |
| rs6456156 | 2.97E-07 | -12.1807 | 4.24E-05 | 0.003482 |
| rs1351030 | 3.20E-07 | -12.1737 | 3.27E-05 | 0.003749 |
| rs2285399 | 3.28E-07 | 12.89861 | 4.60E-05 | 0.003843 |
| rs1051367 | 3.29E-07 | -13.4689 | 7.56E-05 | 0.003864 |
| rs4510072 | 3.30E-07 | -11.8509 | 6.19E-05 | 0.003866 |
| rs1010285 | 3.38E-07 | -13.4383 | 2.85E-05 | 0.003968 |
| rs2110376 | 3.50E-07 | -11.7794 | 4.36E-05 | 0.0041 |
| rs10225945 | 3.53E-07 | -14.2327 | 5.51E-06 | 0.004142 |
| rs2586204 | 3.54E-07 | -11.863 | 5.44E-06 | 0.004148 |
| rs13011264 | 3.55E-07 | -11.7852 | 1.74E-05 | 0.004168 |
| rs7333011 | 3.56E-07 | 12.10822 | 4.67E-05 | 0.004177 |
| rs7601 | 3.57E-07 | 11.74742 | 7.02E-05 | 0.004185 |
| rs4463912 | 3.67E-07 | 13.92157 | 5.06E-05 | 0.004305 |
| rs590329 | 3.69E-07 | -11.9024 | 3.24E-05 | 0.004333 |
| rs113306994 | 3.70E-07 | 13.77595 | 2.80E-05 | 0.004337 |
| rs12428172 | 3.75E-07 | 11.82775 | 3.46E-05 | 0.0044 |
| rs10894266 | 3.80E-07 | 14.8876 | 3.01E-05 | 0.004463 |
| rs1332010 | 3.87E-07 | 12.66734 | 2.81E-05 | 0.004534 |
| rs10838877 | 4.04E-07 | -11.5961 | 1.06E-05 | 0.004741 |
| rs12210295 | 4.10E-07 | 12.6026 | 1.27E-05 | 0.004806 |
| rs7760 | 4.11E-07 | 11.57416 | 9.12E-06 | 0.004826 |
| rs12635996 | 4.19E-07 | -11.573 | 2.03E-05 | 0.004916 |
| rs16887405 | 4.36E-07 | 13.99249 | 7.02E-05 | 0.005116 |
| rs1292336 | 4.39E-07 | 11.53678 | 5.40E-05 | 0.005154 |
| rs35819739 | 4.54E-07 | -11.4518 | 3.14E-06 | 0.005326 |
| rs11204064 | 4.63E-07 | 12.64823 | 1.21E-06 | 0.00543 |
| rs2914004 | 4.72E-07 | 11.42703 | 1.20E-05 | 0.005535 |
| rs436064 | 4.72E-07 | 11.63255 | 9.27E-06 | 0.005536 |
| rs4686952 | 4.72E-07 | -12.1361 | 8.26E-06 | 0.005538 |
| rs2817357 | 4.72E-07 | -11.74 | 2.37E-05 | 0.00554 |
| rs2229313 | 4.85E-07 | -12.0313 | 1.37E-05 | 0.005693 |
| rs3743860 | 4.94E-07 | -11.3748 | 1.44E-05 | 0.005796 |
| rs4740383 | 4.95E-07 | 11.87027 | 4.20E-05 | 0.005807 |
| rs1132780 | 5.04E-07 | 11.76923 | 2.11E-05 | 0.005915 |
| rs9471632 | 5.06E-07 | -11.4288 | 3.20E-06 | 0.005932 |
| rs9640168 | 5.07E-07 | -11.9776 | 0.00011 | 0.005948 |
| rs7915813 | 5.15E-07 | 11.30463 | 1.20E-05 | 0.006041 |
| rs2979082 | 5.25E-07 | -11.2866 | 0.000117 | 0.006153 |
| rs1955915 | 5.36E-07 | 12.12536 | 1.96E-05 | 0.006289 |
| rs56155004 | 5.39E-07 | 12.75579 | 6.21E-05 | 0.006322 |
| rs6125920 | 5.54E-07 | -12.3811 | 4.62E-05 | 0.006494 |
| rs11924254 | 5.55E-07 | 11.5962 | 3.51E-05 | 0.006506 |
| rs6496741 | 5.64E-07 | 11.20124 | 4.44E-06 | 0.006619 |
| rs6484487 | 5.66E-07 | 11.64379 | 4.53E-05 | 0.006633 |
| rs12330479 | 5.76E-07 | 12.83693 | 3.95E-05 | 0.006752 |
| rs10511591 | 5.76E-07 | 11.20884 | 3.33E-05 | 0.006754 |
| rs11228613 | 5.97E-07 | -11.1203 | 3.40E-05 | 0.006997 |
| rs2583949 | 6.07E-07 | 11.56241 | 0.000125 | 0.007122 |
| rs4074448 | 6.10E-07 | 12.20874 | 4.64E-05 | 0.007153 |
| rs1043654 | 6.17E-07 | 11.5715 | 2.12E-05 | 0.007234 |
| rs60637 | 6.19E-07 | 11.1765 | 6.22E-05 | 0.007259 |
| rs9923967 | 6.29E-07 | -11.2317 | 8.03E-06 | 0.00738 |
| rs12880628 | 6.86E-07 | 11.56612 | 1.09E-05 | 0.008045 |
| rs56134233 | 6.87E-07 | -11.2103 | 3.12E-05 | 0.008053 |
| rs1978982 | 6.98E-07 | -11.7233 | 6.62E-06 | 0.00819 |
| rs3129287 | 7.10E-07 | 11.56691 | 6.65E-06 | 0.008325 |
| rs6993025 | 7.14E-07 | 11.26768 | 8.85E-05 | 0.008371 |
| rs11509880 | 7.16E-07 | 11.2818 | 6.32E-05 | 0.0084 |
| rs61856637 | 7.21E-07 | -10.908 | 5.11E-05 | 0.008454 |
| rs57585717 | 7.21E-07 | -11.2503 | 2.76E-05 | 0.008457 |
| rs4767649 | 7.26E-07 | -10.8892 | 1.43E-05 | 0.008514 |
| rs3960788 | 7.26E-07 | -11.1443 | 3.46E-05 | 0.008514 |
| rs10930140 | 7.28E-07 | 11.08111 | 5.46E-05 | 0.008544 |
| rs1709852 | 7.33E-07 | 11.70671 | 4.40E-05 | 0.008593 |
| rs723490 | 7.36E-07 | -10.9935 | 2.24E-05 | 0.008633 |
| rs2320225 | 7.42E-07 | 10.87091 | 3.93E-05 | 0.008703 |
| rs4724449 | 7.45E-07 | 11.48955 | 4.19E-05 | 0.008737 |
| rs704447 | 7.56E-07 | 11.08684 | 3.45E-06 | 0.008866 |
| rs2927305 | 7.66E-07 | 14.27038 | 6.96E-05 | 0.008986 |
| rs740123 | 7.67E-07 | 11.39719 | 7.89E-05 | 0.008992 |
| rs858519 | 7.69E-07 | 11.12269 | 5.24E-05 | 0.00902 |
| rs2838813 | 7.77E-07 | -15.1588 | 6.01E-05 | 0.009112 |
| rs6592775 | 7.84E-07 | 12.01677 | 1.95E-05 | 0.009199 |
| rs3094221 | 7.90E-07 | 12.46838 | 2.99E-05 | 0.009261 |
| rs55917610 | 7.93E-07 | -10.9361 | 1.18E-05 | 0.009297 |
| rs1893562 | 7.94E-07 | -10.9048 | 7.27E-06 | 0.009311 |
| rs3791696 | 8.06E-07 | -10.7735 | 9.93E-06 | 0.009457 |
| rs6982912 | 8.23E-07 | -11.1843 | 9.62E-07 | 0.009654 |
| rs2856997 | 8.39E-07 | 11.22648 | 4.20E-05 | 0.009844 |
| rs12879453 | 8.50E-07 | -10.7461 | 1.83E-05 | 0.00997 |
| rs6565066 | 8.71E-07 | -10.6859 | 4.34E-05 | 0.010216 |
| rs2922979 | 9.05E-07 | -10.99 | 4.78E-05 | 0.010621 |
| rs12461274 | 9.23E-07 | 11.81577 | 4.07E-05 | 0.010832 |
| rs3818717 | 9.38E-07 | 11.60007 | 7.66E-06 | 0.010998 |
| rs1871900 | 9.47E-07 | -10.6647 | 2.45E-06 | 0.011106 |
| rs10770662 | 9.47E-07 | -10.6718 | 8.25E-06 | 0.01111 |
| rs12809441 | 9.57E-07 | -10.6802 | 3.65E-05 | 0.011231 |
| rs2238448 | 9.81E-07 | 12.01907 | 1.99E-06 | 0.011511 |
| rs12545240 | 1.00E-06 | 12.02419 | 0.000104 | 0.011747 |
| rs11041830 | 1.01E-06 | -10.5979 | 1.21E-06 | 0.011905 |
| rs10742277 | 1.02E-06 | 14.07352 | 2.82E-05 | 0.011921 |
| rs2253569 | 1.04E-06 | -12.7545 | 1.10E-05 | 0.012141 |
| rs1861869 | 1.06E-06 | 10.91951 | 5.07E-05 | 0.012466 |
| rs61748245 | 1.07E-06 | -10.54 | 2.26E-05 | 0.012589 |
| rs7030475 | 1.08E-06 | 10.44526 | 7.99E-05 | 0.012687 |
| rs1402837 | 1.10E-06 | -10.4798 | 8.18E-06 | 0.012854 |
| rs5752783 | 1.10E-06 | 10.4321 | 5.57E-05 | 0.012943 |
| rs61572685 | 1.10E-06 | -10.4878 | 1.16E-05 | 0.012953 |
| rs2665303 | 1.11E-06 | 10.44027 | 4.31E-05 | 0.012998 |
| rs7095472 | 1.11E-06 | 10.52328 | 1.97E-05 | 0.013046 |
| rs11645687 | 1.11E-06 | 10.44835 | 2.08E-05 | 0.013056 |
| rs11031783 | 1.12E-06 | 11.41561 | 2.44E-05 | 0.013133 |
| rs10937048 | 1.13E-06 | 10.89896 | 4.24E-05 | 0.01322 |
| rs4513466 | 1.15E-06 | -10.6123 | 2.70E-05 | 0.013447 |
| rs6444191 | 1.15E-06 | -10.4361 | 3.08E-06 | 0.013536 |
| rs1267673 | 1.16E-06 | -11.1245 | 2.25E-05 | 0.01362 |
| rs9825233 | 1.16E-06 | 10.35344 | 1.92E-05 | 0.013629 |
| rs6444082 | 1.17E-06 | 10.36767 | 0.000133 | 0.013695 |
| rs1895874 | 1.17E-06 | -10.348 | 1.65E-05 | 0.013745 |
| rs2025776 | 1.18E-06 | 10.59082 | 1.92E-05 | 0.013783 |
| rs6927207 | 1.18E-06 | 12.4717 | 1.16E-05 | 0.013836 |
| rs68024867 | 1.18E-06 | -10.3274 | 3.51E-05 | 0.013885 |
| rs1906397 | 1.19E-06 | -10.3349 | 1.40E-05 | 0.013946 |
| rs1139266 | 1.19E-06 | -11.208 | 3.89E-05 | 0.014003 |
| rs818382 | 1.20E-06 | -10.77 | 1.11E-05 | 0.014059 |
| rs4150944 | 1.20E-06 | -14.3674 | 8.12E-06 | 0.01408 |
| rs12608943 | 1.20E-06 | 10.48762 | 3.38E-05 | 0.014095 |
| rs72874866 | 1.27E-06 | 11.05813 | 8.71E-05 | 0.014881 |
| rs6444790 | 1.28E-06 | -10.4943 | 5.39E-05 | 0.015035 |
| rs892609 | 1.30E-06 | -10.3076 | 2.58E-05 | 0.015214 |
| rs764117 | 1.30E-06 | -10.7282 | 4.11E-05 | 0.01522 |
| rs11040535 | 1.33E-06 | 10.95066 | 1.84E-05 | 0.015657 |
| rs1893174 | 1.35E-06 | 10.36604 | 1.42E-05 | 0.015797 |
| rs56239097 | 1.37E-06 | -10.4079 | 5.10E-05 | 0.016073 |
| rs2186120 | 1.39E-06 | -10.6351 | 1.67E-05 | 0.016322 |
| rs13277568 | 1.40E-06 | -11.0569 | 0.000115 | 0.016418 |
| rs773410 | 1.40E-06 | 10.71924 | 5.40E-05 | 0.016421 |
| rs11726779 | 1.41E-06 | -10.1456 | 6.52E-06 | 0.016586 |
| rs6809679 | 1.42E-06 | -10.1251 | 7.47E-05 | 0.016707 |
| rs373857 | 1.43E-06 | 16.7897 | 2.08E-05 | 0.016723 |
| rs653170 | 1.44E-06 | -10.145 | 2.13E-05 | 0.016835 |
| rs1406755 | 1.46E-06 | -10.5136 | 5.49E-06 | 0.017075 |
| rs3848572 | 1.46E-06 | 10.21504 | 2.90E-05 | 0.017107 |
| rs7533344 | 1.47E-06 | -10.8496 | 5.93E-07 | 0.01729 |
| rs1405630 | 1.51E-06 | 10.29785 | 5.80E-05 | 0.017657 |
| rs1250560 | 1.52E-06 | 10.05809 | 5.63E-06 | 0.017851 |
| rs1900272 | 1.54E-06 | 10.38016 | 2.94E-06 | 0.018019 |
| rs13018340 | 1.54E-06 | -10.9832 | 2.17E-06 | 0.018078 |
| rs2506141 | 1.54E-06 | -11.2444 | 4.11E-05 | 0.018107 |
| rs7023690 | 1.54E-06 | 10.09068 | 1.87E-06 | 0.018116 |
| rs12450045 | 1.55E-06 | 12.99343 | 5.95E-05 | 0.018212 |
| rs6545144 | 1.56E-06 | 11.00754 | 2.56E-05 | 0.018258 |
| rs12995245 | 1.57E-06 | 10.1102 | 2.17E-05 | 0.018384 |
| rs10131172 | 1.57E-06 | 10.01298 | 1.93E-05 | 0.018432 |
| rs2546110 | 1.58E-06 | -10.2341 | 1.45E-05 | 0.018479 |
| rs2714309 | 1.59E-06 | 10.00126 | 2.39E-05 | 0.018681 |
| rs10937778 | 1.59E-06 | 10.65032 | 7.21E-05 | 0.018698 |
| rs1859572 | 1.59E-06 | 10.0435 | 4.92E-05 | 0.018699 |
| rs1488119 | 1.62E-06 | -10.8094 | 1.82E-05 | 0.01905 |
| rs1971396 | 1.64E-06 | 14.0777 | 4.18E-05 | 0.01925 |
| rs1993376 | 1.65E-06 | -10.7788 | 3.18E-05 | 0.019356 |
| rs12885354 | 1.66E-06 | -10.6122 | 4.36E-05 | 0.019505 |
| rs10804881 | 1.69E-06 | -9.93667 | 7.14E-05 | 0.019782 |
| rs7425339 | 1.69E-06 | -10.2371 | 2.96E-05 | 0.019833 |
| rs28818616 | 1.71E-06 | -10.1237 | 2.40E-05 | 0.020032 |
| rs10108060 | 1.71E-06 | -10.0256 | 2.15E-06 | 0.020041 |
| rs12636004 | 1.73E-06 | 10.11672 | 2.98E-05 | 0.02031 |
| rs1641549 | 1.73E-06 | 10.14795 | 2.81E-05 | 0.020351 |
| rs2842873 | 1.74E-06 | -10.1 | 2.98E-05 | 0.020387 |
| rs7789785 | 1.75E-06 | 9.947042 | 3.58E-05 | 0.020561 |
| rs519856 | 1.75E-06 | -13.0992 | 6.18E-06 | 0.020575 |
| rs7206010 | 1.77E-06 | -14.826 | 2.83E-05 | 0.020725 |
| rs2291027 | 1.78E-06 | 12.90356 | 2.71E-05 | 0.020896 |
| rs2108635 | 1.79E-06 | 10.02542 | 4.56E-05 | 0.020946 |
| rs7254282 | 1.81E-06 | 13.73662 | 2.48E-05 | 0.021246 |
| rs13387347 | 1.81E-06 | 9.871811 | 1.99E-05 | 0.021278 |
| rs657693 | 1.82E-06 | 9.856958 | 2.16E-05 | 0.021336 |
| rs16850985 | 1.82E-06 | 10.84533 | 2.83E-05 | 0.021337 |
| rs7219571 | 1.83E-06 | -9.93186 | 8.23E-06 | 0.021415 |
| rs17834412 | 1.84E-06 | -9.90684 | 5.36E-05 | 0.021545 |
| rs1622555 | 1.84E-06 | -11.5262 | 5.38E-05 | 0.021557 |
| rs16955396 | 1.85E-06 | -10.2915 | 6.63E-06 | 0.021698 |
| rs4234677 | 1.87E-06 | 10.13862 | 2.61E-05 | 0.021898 |
| rs35845603 | 1.87E-06 | 10.94421 | 7.84E-05 | 0.021922 |
| rs35338539 | 1.88E-06 | 10.42096 | 6.76E-05 | 0.022081 |
| rs6977416 | 1.88E-06 | -9.9303 | 1.45E-05 | 0.022106 |
| rs1836718 | 1.89E-06 | 13.78124 | 1.23E-05 | 0.022114 |
| rs2703805 | 1.89E-06 | -10.4509 | 4.28E-05 | 0.022154 |
| rs763073 | 1.92E-06 | 10.77649 | 3.42E-05 | 0.022522 |
| rs10768108 | 1.93E-06 | 9.794962 | 2.31E-05 | 0.022586 |
| rs3131003 | 1.95E-06 | 10.21441 | 6.17E-05 | 0.02288 |
| rs9290404 | 1.97E-06 | -9.86686 | 1.34E-05 | 0.023091 |
| rs6466626 | 1.97E-06 | 10.61039 | 2.33E-05 | 0.023106 |
| rs3130453 | 1.98E-06 | 13.7287 | 1.59E-05 | 0.023241 |
| rs9502637 | 1.99E-06 | -10.2823 | 2.53E-05 | 0.023324 |
| rs2059249 | 2.00E-06 | -10.319 | 9.49E-05 | 0.023493 |
| rs6480771 | 2.01E-06 | 9.748256 | 4.05E-05 | 0.023588 |
| rs12376677 | 2.05E-06 | -9.80357 | 3.57E-05 | 0.024069 |
| rs7526262 | 2.06E-06 | 9.762304 | 1.45E-05 | 0.024121 |
| rs16846133 | 2.06E-06 | -11.2328 | 2.89E-05 | 0.024129 |
| rs10499183 | 2.06E-06 | 12.20082 | 1.56E-05 | 0.024143 |
| rs12712919 | 2.07E-06 | -10.0788 | 2.04E-05 | 0.024315 |
| rs17221059 | 2.07E-06 | 10.12417 | 1.05E-05 | 0.024334 |
| rs17563158 | 2.09E-06 | 9.730409 | 8.27E-05 | 0.024564 |
| rs4456287 | 2.15E-06 | -9.6774 | 8.77E-05 | 0.025189 |
| rs7743221 | 2.16E-06 | 10.18674 | 3.34E-05 | 0.025288 |
| rs2504228 | 2.17E-06 | 10.60627 | 6.37E-05 | 0.025429 |
| rs6962676 | 2.20E-06 | -10.5319 | 3.79E-05 | 0.025776 |
| rs1989208 | 2.21E-06 | -9.66111 | 8.82E-06 | 0.025919 |
| rs3791682 | 2.21E-06 | -10.0502 | 2.51E-05 | 0.025967 |
| rs12912147 | 2.22E-06 | 9.902935 | 3.47E-05 | 0.026003 |
| rs4436026 | 2.22E-06 | -10.8225 | 1.03E-05 | 0.026027 |
| rs4923889 | 2.22E-06 | 13.82138 | 8.14E-06 | 0.026039 |
| rs2255750 | 2.24E-06 | 10.17546 | 3.58E-05 | 0.026231 |
| rs7023663 | 2.25E-06 | 9.660535 | 3.42E-05 | 0.026355 |
| rs10774905 | 2.25E-06 | 10.31102 | 8.33E-07 | 0.02638 |
| rs7188755 | 2.28E-06 | -10.3256 | 2.28E-05 | 0.02673 |
| rs8057014 | 2.36E-06 | -9.60764 | 1.69E-05 | 0.027722 |
| rs1025912 | 2.37E-06 | -10.056 | 1.57E-05 | 0.027797 |
| rs11190270 | 2.39E-06 | -9.59002 | 1.92E-05 | 0.028079 |
| rs10245867 | 2.42E-06 | 11.01204 | 9.06E-05 | 0.028413 |
| rs9788998 | 2.49E-06 | -9.93352 | 4.19E-05 | 0.029256 |
| rs1965788 | 2.50E-06 | -11.6726 | 2.05E-06 | 0.029269 |
| rs10209654 | 2.52E-06 | -9.56164 | 7.27E-06 | 0.029601 |
| rs1020526 | 2.55E-06 | -10.363 | 7.27E-06 | 0.029968 |
| rs4620747 | 2.57E-06 | -9.64121 | 2.69E-05 | 0.030105 |
| rs1082034 | 2.58E-06 | -9.69347 | 6.73E-05 | 0.030213 |
| rs7758790 | 2.59E-06 | 9.485623 | 4.37E-05 | 0.030331 |
| rs10772620 | 2.61E-06 | 9.696416 | 5.73E-06 | 0.030648 |
| rs3006070 | 2.62E-06 | 9.495839 | 3.42E-05 | 0.030699 |
| rs4794758 | 2.62E-06 | 11.07588 | 3.39E-06 | 0.030744 |
| rs840969 | 2.63E-06 | 9.479407 | 4.39E-05 | 0.030883 |
| rs2179591 | 2.64E-06 | 9.49271 | 6.60E-05 | 0.030954 |
| rs936598 | 2.65E-06 | -9.48751 | 4.76E-05 | 0.03106 |
| rs12949853 | 2.65E-06 | 11.23341 | 4.25E-06 | 0.031094 |
| rs10280605 | 2.65E-06 | 9.723509 | 2.85E-06 | 0.031095 |
| rs10172668 | 2.66E-06 | -10.7277 | 1.65E-05 | 0.031198 |
| rs11084922 | 2.66E-06 | -10.1214 | 0.000111 | 0.031244 |
| rs9851055 | 2.67E-06 | -12.2721 | 1.10E-05 | 0.031291 |
| rs12614265 | 2.69E-06 | -9.46957 | 4.67E-05 | 0.031524 |
| rs9405331 | 2.69E-06 | 13.00374 | 2.85E-05 | 0.03153 |
| rs9897114 | 2.71E-06 | -9.4589 | 8.80E-06 | 0.031737 |
| rs4933220 | 2.73E-06 | -10.593 | 7.99E-05 | 0.032019 |
| rs1637868 | 2.75E-06 | -9.8047 | 1.40E-05 | 0.032203 |
| rs178845 | 2.77E-06 | 10.58021 | 2.44E-05 | 0.032448 |
| rs185963 | 2.78E-06 | 10.34968 | 8.71E-05 | 0.032617 |
| rs7193297 | 2.80E-06 | -9.60957 | 8.98E-06 | 0.032841 |
| rs9442200 | 2.81E-06 | 9.924663 | 7.89E-06 | 0.032922 |
| rs11082685 | 2.83E-06 | -9.82323 | 8.96E-05 | 0.033239 |
| rs6128408 | 2.85E-06 | 9.803384 | 1.42E-05 | 0.033455 |
| rs6979947 | 2.96E-06 | -10.5088 | 7.21E-05 | 0.034729 |
| rs3116095 | 2.98E-06 | -9.74856 | 3.01E-06 | 0.035002 |
| rs13259464 | 3.02E-06 | -9.64007 | 2.95E-05 | 0.035382 |
| rs10734253 | 3.04E-06 | -9.45296 | 6.12E-05 | 0.035677 |
| rs3826655 | 3.04E-06 | 10.4647 | 4.38E-05 | 0.035693 |
| rs1610645 | 3.11E-06 | 10.39676 | 1.50E-05 | 0.036456 |
| rs7871600 | 3.11E-06 | -9.52422 | 3.40E-05 | 0.036476 |
| rs1250552 | 3.14E-06 | 10.04939 | 8.48E-05 | 0.036859 |
| rs10973541 | 3.14E-06 | -9.32802 | 6.09E-05 | 0.036861 |
| rs666194 | 3.14E-06 | 9.536225 | 5.23E-05 | 0.036865 |
| rs9513018 | 3.14E-06 | 9.286337 | 8.31E-05 | 0.036889 |
| rs2252641 | 3.16E-06 | -9.44715 | 2.44E-05 | 0.037019 |
| rs7341574 | 3.18E-06 | -11.0858 | 2.30E-07 | 0.037341 |
| rs17855765 | 3.20E-06 | 9.595034 | 5.33E-06 | 0.037591 |
| rs1291324 | 3.23E-06 | 10.95795 | 5.97E-06 | 0.037865 |
| rs12959719 | 3.25E-06 | -10.0507 | 2.03E-05 | 0.038109 |
| rs62383436 | 3.26E-06 | 9.836054 | 0.000129 | 0.038196 |
| rs1085485 | 3.26E-06 | -9.36772 | 4.43E-05 | 0.038249 |
| rs2568465 | 3.29E-06 | 9.439238 | 4.31E-06 | 0.038581 |
| rs11941852 | 3.30E-06 | 11.55651 | 5.67E-05 | 0.038722 |
| rs11264357 | 3.32E-06 | 9.310281 | 4.04E-05 | 0.038944 |
| rs9990527 | 3.32E-06 | -9.40628 | 3.88E-06 | 0.038951 |
| rs350094 | 3.34E-06 | 9.245072 | 1.28E-05 | 0.039134 |
| rs2298999 | 3.36E-06 | 9.265663 | 4.22E-05 | 0.039406 |
| rs34341 | 3.36E-06 | -12.0853 | 0.000103 | 0.039453 |
| rs8791 | 3.37E-06 | 12.45165 | 3.24E-05 | 0.039557 |
| rs10228495 | 3.38E-06 | -9.29063 | 6.87E-05 | 0.039621 |
| rs2112653 | 3.39E-06 | 11.16177 | 5.41E-06 | 0.039723 |
| rs168144 | 3.41E-06 | -10.1643 | 3.20E-05 | 0.039944 |
| rs9527175 | 3.42E-06 | 9.348168 | 2.24E-05 | 0.040125 |
| rs11128523 | 3.44E-06 | -9.18536 | 3.37E-05 | 0.040397 |
| rs7147370 | 3.44E-06 | 9.185265 | 2.74E-05 | 0.040407 |
| rs39916 | 3.52E-06 | 9.394491 | 3.59E-05 | 0.041296 |
| rs6722613 | 3.54E-06 | 9.856644 | 8.72E-05 | 0.041522 |
| rs4712536 | 3.57E-06 | 10.00063 | 2.78E-06 | 0.041841 |
| rs4793090 | 3.59E-06 | 9.168978 | 5.83E-05 | 0.04214 |
| rs7975539 | 3.63E-06 | -9.15144 | 2.92E-06 | 0.042535 |
| rs2396369 | 3.69E-06 | 10.11272 | 2.45E-05 | 0.043235 |
| rs6972713 | 3.73E-06 | 11.11921 | 3.24E-05 | 0.043708 |
| rs2864434 | 3.75E-06 | 9.520515 | 5.29E-06 | 0.043992 |
| rs13000621 | 3.78E-06 | 9.093298 | 4.18E-05 | 0.044345 |
| rs2127162 | 3.81E-06 | -9.2522 | 6.47E-05 | 0.044665 |
| rs6963810 | 3.81E-06 | -9.42551 | 9.06E-07 | 0.044686 |
| rs10851925 | 3.81E-06 | -9.16594 | 2.67E-05 | 0.04473 |
| rs806187 | 3.81E-06 | -10.8589 | 4.53E-05 | 0.044745 |
| rs2250931 | 3.82E-06 | -11.3816 | 1.86E-06 | 0.044861 |
| rs1915241 | 3.85E-06 | -10.8084 | 7.71E-05 | 0.045164 |
| rs460214 | 3.89E-06 | 9.09825 | 3.99E-05 | 0.045676 |
| rs1941141 | 3.92E-06 | 12.24751 | 2.56E-05 | 0.046012 |
| rs1291822 | 3.95E-06 | -9.07293 | 1.88E-05 | 0.046379 |
| rs12417381 | 3.96E-06 | 9.150853 | 1.23E-05 | 0.046453 |
| rs5770890 | 3.96E-06 | 9.294668 | 4.94E-05 | 0.046478 |
| rs13248612 | 4.05E-06 | 11.19865 | 3.18E-05 | 0.047474 |
| rs1076849 | 4.06E-06 | 9.094962 | 6.92E-05 | 0.047574 |
| rs4692523 | 4.13E-06 | 9.970252 | 2.10E-05 | 0.048409 |
| rs12485473 | 4.14E-06 | -9.06629 | 1.92E-05 | 0.048603 |
| rs3773691 | 4.15E-06 | 11.18388 | 4.93E-05 | 0.048641 |
| rs6892212 | 4.17E-06 | 8.992768 | 1.84E-05 | 0.048899 |
| rs12934833 | 4.21E-06 | 10.86993 | 3.25E-05 | 0.049342 |
| rs72747464 | 4.21E-06 | 9.128069 | 6.66E-05 | 0.049348 |
| rs9358383 | 4.26E-06 | -9.08789 | 1.79E-05 | 0.049939 |

**Supplementary Table 2: CID-T2D Adv-PC Model Feature Importance T-test Results**

| **SNP** | **t_test_p_value** | **t_test_statistic** | **t_test_std_error** | **adjusted_p_value** |
| --- | --- | --- | --- | --- |
| rs73235142 | 1.84E-09 | 20.7976 | 1.52E-06 | 2.16E-05 |
| rs138771 | 4.39E-08 | 15.03097 | 1.86E-06 | 0.000515 |
| rs1004531 | 5.86E-08 | 14.71772 | 2.63E-06 | 0.000687 |
| rs935376 | 6.35E-08 | 14.17789 | 4.08E-06 | 0.000745 |
| rs9825233 | 6.95E-08 | 15.43571 | 4.90E-06 | 0.000815 |
| rs12330479 | 7.49E-08 | 14.07741 | 2.21E-06 | 0.000879 |
| rs4258685 | 1.03E-07 | 14.11876 | 3.58E-06 | 0.001209 |
| rs9897114 | 1.36E-07 | 13.26854 | 7.96E-06 | 0.001592 |
| rs6504663 | 2.61E-07 | -12.4838 | 2.09E-06 | 0.003066 |
| rs1020526 | 2.91E-07 | 12.40313 | 6.91E-06 | 0.003414 |
| rs4299668 | 3.22E-07 | 12.47497 | 4.23E-06 | 0.003778 |
| rs4234677 | 3.84E-07 | -13.3762 | 2.31E-06 | 0.004505 |
| rs165599 | 3.97E-07 | 11.86334 | 4.84E-06 | 0.004663 |
| rs13256092 | 4.14E-07 | -12.2353 | 2.57E-06 | 0.004853 |
| rs6444191 | 4.29E-07 | -13.2317 | 5.17E-06 | 0.005028 |
| rs9680643 | 4.69E-07 | 11.48083 | 1.79E-06 | 0.005503 |
| rs6764532 | 5.41E-07 | 11.45939 | 4.30E-06 | 0.006348 |
| rs2927305 | 5.96E-07 | 11.60211 | 3.51E-06 | 0.006993 |
| rs1260327 | 6.24E-07 | 11.15725 | 1.05E-05 | 0.007317 |
| rs4793090 | 7.24E-07 | 12.7157 | 5.44E-06 | 0.008492 |
| rs7873848 | 8.16E-07 | -10.9574 | 4.14E-06 | 0.009566 |
| rs1192924 | 8.45E-07 | 10.79072 | 3.01E-06 | 0.009917 |
| rs3732675 | 8.92E-07 | 11.0545 | 5.45E-06 | 0.010463 |
| rs6503052 | 9.11E-07 | 11.05574 | 5.73E-06 | 0.010683 |
| rs7219571 | 9.34E-07 | 10.62484 | 2.75E-06 | 0.010952 |
| rs1982151 | 1.02E-06 | 10.4937 | 5.81E-06 | 0.01197 |
| rs2409717 | 1.06E-06 | -12.966 | 3.07E-06 | 0.012403 |
| rs12635996 | 1.27E-06 | 11.08895 | 3.29E-06 | 0.01492 |
| rs9395049 | 1.51E-06 | 11.11572 | 7.87E-06 | 0.017679 |
| rs4074683 | 1.52E-06 | 10.1166 | 1.82E-06 | 0.017827 |
| rs11517737 | 1.71E-06 | -9.95309 | 2.98E-06 | 0.020056 |
| rs4929992 | 1.72E-06 | -9.97595 | 5.50E-06 | 0.020218 |
| rs3794550 | 1.88E-06 | -11.3901 | 2.22E-06 | 0.022036 |
| rs7640 | 1.90E-06 | 11.40008 | 4.51E-06 | 0.022342 |
| rs2134279 | 1.95E-06 | 11.03152 | 1.64E-06 | 0.022905 |
| rs5762936 | 2.01E-06 | 12.17293 | 3.17E-06 | 0.02353 |
| rs9866303 | 2.12E-06 | -9.78535 | 2.71E-06 | 0.024901 |
| rs1157350 | 2.15E-06 | 9.758774 | 6.59E-06 | 0.025174 |
| rs72717965 | 2.17E-06 | 9.758631 | 2.31E-06 | 0.025418 |
| rs4074448 | 2.31E-06 | 10.19293 | 6.80E-06 | 0.027139 |
| rs2583930 | 2.34E-06 | 9.609082 | 3.04E-05 | 0.027403 |
| rs2071473 | 2.51E-06 | 9.618591 | 7.06E-06 | 0.02949 |
| rs13122948 | 2.55E-06 | -10.0405 | 3.39E-06 | 0.029957 |
| rs2842353 | 2.68E-06 | 12.51271 | 5.74E-06 | 0.031423 |
| rs1275195 | 2.71E-06 | 13.20376 | 4.43E-06 | 0.031769 |
| rs2583949 | 2.72E-06 | 10.94692 | 7.61E-06 | 0.031876 |
| rs1325195 | 2.82E-06 | 12.35781 | 3.93E-06 | 0.033106 |
| rs395987 | 3.17E-06 | 9.657763 | 6.23E-06 | 0.037201 |
| rs1213769 | 3.24E-06 | -10.3315 | 2.40E-06 | 0.038002 |
| rs17025712 | 3.35E-06 | -11.0323 | 2.10E-06 | 0.039261 |
| rs1776196 | 3.46E-06 | -11.4291 | 3.25E-06 | 0.040531 |
| rs2839196 | 3.63E-06 | 9.15427 | 6.95E-06 | 0.04263 |
| rs763073 | 3.73E-06 | 9.356648 | 9.41E-06 | 0.043751 |
| rs4463912 | 3.76E-06 | 9.638827 | 5.06E-06 | 0.044154 |
| rs7796576 | 3.89E-06 | 9.383576 | 4.35E-06 | 0.045642 |
| rs1941141 | 4.00E-06 | 9.621209 | 3.18E-06 | 0.046875 |
| rs6692491 | 4.10E-06 | -9.10305 | 1.59E-06 | 0.048076 |

**Supplementary Table 3: CID Specific Model Feature Importance T-test Results**

| **SNP** | **t_test_p_value** | **t_test_statistic** | **t_test_std_error** | **adjusted_p_value** |
| --- | --- | --- | --- | --- |
| rs1267673 | 1.49E-08 | 16.89143 | 7.01E-06 | 0.000175 |
| rs7917983 | 2.57E-08 | 15.57126 | 1.09E-05 | 0.000301 |
| rs7225149 | 3.66E-08 | 15.37261 | 4.11E-06 | 0.00043 |
| rs9289008 | 6.84E-08 | -16.8955 | 7.36E-06 | 0.000803 |
| rs823155 | 7.46E-08 | -15.0607 | 9.83E-06 | 0.000875 |
| rs1941141 | 8.22E-08 | 13.71957 | 5.42E-06 | 0.000964 |
| rs17115870 | 9.65E-08 | 13.73321 | 8.88E-06 | 0.001132 |
| rs17049683 | 1.17E-07 | -13.5065 | 4.72E-06 | 0.001367 |
| rs7672622 | 2.20E-07 | 12.36863 | 1.10E-05 | 0.002582 |
| rs4505038 | 2.37E-07 | 16.67026 | 1.13E-05 | 0.002786 |
| rs7855329 | 2.91E-07 | 12.01146 | 1.26E-05 | 0.003419 |
| rs1151572 | 3.47E-07 | 11.99935 | 3.88E-06 | 0.004076 |
| rs11000785 | 4.22E-07 | 14.11493 | 1.29E-05 | 0.004956 |
| rs2981422 | 4.24E-07 | -11.6318 | 1.26E-05 | 0.004979 |
| rs11985674 | 5.07E-07 | -11.4035 | 6.66E-06 | 0.005943 |
| rs5218 | 5.12E-07 | 13.84584 | 9.32E-06 | 0.00601 |
| rs17221059 | 5.72E-07 | 11.21119 | 5.06E-06 | 0.006713 |
| rs7025006 | 6.03E-07 | -11.4351 | 8.14E-06 | 0.007079 |
| rs7166725 | 6.13E-07 | 11.29114 | 6.61E-06 | 0.00719 |
| rs243527 | 6.22E-07 | 15.41629 | 1.02E-05 | 0.0073 |
| rs11097249 | 6.78E-07 | 15.76361 | 9.57E-06 | 0.007951 |
| rs3757840 | 6.87E-07 | -11.2735 | 2.04E-05 | 0.008054 |
| rs3791710 | 6.97E-07 | 11.05594 | 5.02E-06 | 0.008172 |
| rs4445867 | 7.19E-07 | 10.89742 | 1.12E-05 | 0.008436 |
| rs4435039 | 8.08E-07 | 10.83569 | 1.37E-05 | 0.00948 |
| rs1836718 | 8.51E-07 | 12.99364 | 7.99E-06 | 0.00998 |
| rs4767903 | 8.84E-07 | 11.11535 | 1.35E-05 | 0.010372 |
| rs62180403 | 1.10E-06 | -10.4224 | 7.10E-06 | 0.012848 |
| rs1494092 | 1.12E-06 | 10.68112 | 1.04E-05 | 0.013154 |
| rs2214884 | 1.13E-06 | 10.47471 | 1.77E-05 | 0.013297 |
| rs6444188 | 1.15E-06 | 10.36113 | 7.41E-06 | 0.013458 |
| rs4720476 | 1.20E-06 | -11.5615 | 7.09E-06 | 0.014031 |
| rs13150002 | 1.26E-06 | 10.79449 | 1.15E-05 | 0.014799 |
| rs2292749 | 1.31E-06 | -11.1633 | 1.74E-05 | 0.0154 |
| rs7126224 | 1.36E-06 | -10.1956 | 1.28E-05 | 0.016009 |
| rs10234845 | 1.39E-06 | -13.584 | 4.39E-06 | 0.01629 |
| rs9845457 | 1.40E-06 | 13.35334 | 1.07E-05 | 0.01638 |
| rs11070863 | 1.44E-06 | -11.408 | 1.20E-05 | 0.016911 |
| rs11187133 | 1.52E-06 | 10.37558 | 9.73E-06 | 0.01785 |
| rs4259245 | 1.55E-06 | -10.1718 | 1.31E-05 | 0.018209 |
| rs4255018 | 1.78E-06 | -9.97251 | 1.58E-05 | 0.02087 |
| rs6809679 | 1.82E-06 | 10.8609 | 5.50E-06 | 0.02131 |
| rs11719375 | 1.90E-06 | 9.978209 | 8.72E-06 | 0.022321 |
| rs796057 | 1.94E-06 | 10.07458 | 9.97E-06 | 0.022705 |
| rs2303771 | 1.97E-06 | -11.2007 | 6.09E-06 | 0.023096 |
| rs1355782 | 2.29E-06 | -9.98061 | 7.24E-06 | 0.026844 |
| rs3809770 | 2.32E-06 | 9.593795 | 1.61E-05 | 0.027231 |
| rs4850956 | 2.35E-06 | 10.67963 | 8.24E-06 | 0.027558 |
| rs9373367 | 2.76E-06 | 9.679797 | 5.65E-06 | 0.032319 |
| rs12166583 | 2.91E-06 | 9.576896 | 8.65E-06 | 0.034092 |
| rs1052656 | 2.91E-06 | 9.622936 | 9.82E-06 | 0.034133 |
| rs6984206 | 2.95E-06 | -9.9913 | 8.69E-06 | 0.034597 |
| rs1325195 | 3.03E-06 | 10.49463 | 8.30E-06 | 0.035599 |
| rs10774905 | 3.37E-06 | -9.60293 | 1.11E-05 | 0.03954 |
| rs2235808 | 3.43E-06 | -10.9447 | 3.93E-06 | 0.040291 |
| rs13280053 | 3.50E-06 | -9.17418 | 7.12E-06 | 0.041052 |
| rs2926590 | 3.56E-06 | 9.181297 | 7.93E-06 | 0.041786 |
| rs35568293 | 3.60E-06 | -9.74967 | 8.74E-06 | 0.042246 |
| rs34127110 | 3.76E-06 | 9.316659 | 7.30E-06 | 0.044131 |
| rs9289415 | 3.81E-06 | -9.29954 | 5.15E-06 | 0.044685 |
| rs7043040 | 3.87E-06 | -10.1021 | 3.41E-06 | 0.045379 |
| rs2286929 | 4.12E-06 | 9.027559 | 1.30E-05 | 0.048354 |

**Supplementary Table 4: MAGMA Results on Training Subset**

| **FULL_NAME** | **NGENES** | **BETA** | **BETA_STD** | **SE** | **P** |
| --- | --- | --- | --- | --- | --- |
| GO_NEGATIVE_REGULATION_OF_TYPE_B_PANCREATIC_CELL_APOPTOTIC_PROCESS | 6 | 3.1 | 0.057 | 0.57 | 2.79E-08 |
| GO_RESPONSE_TO_CARBOHYDRATE | 213 | 0.427 | 0.0466 | 0.0786 | 2.82E-08 |
| GO_CARBOHYDRATE_HOMEOSTASIS | 223 | 0.416 | 0.0464 | 0.078 | 4.91E-08 |
| GO_FIBROBLAST_GROWTH_FACTOR_ACTIVATED_RECEPTOR_ACTIVITY | 5 | 2.86 | 0.048 | 0.559 | 1.62E-07 |
| GO_POSITIVE_REGULATION_OF_NUCLEOBASE_CONTAINING_COMPOUND_METABOLIC_PROCESS | 1723 | 0.149 | 0.0441 | 0.0296 | 2.60E-07 |
| GO_REGULATION_OF_SEQUESTERING_OF_ZINC_ION | 5 | 2.99 | 0.0502 | 0.603 | 3.68E-07 |
| GO_INSULIN_SECRETION | 194 | 0.414 | 0.0431 | 0.0844 | 4.80E-07 |
| GO_PANCREAS_DEVELOPMENT | 75 | 0.677 | 0.044 | 0.139 | 5.66E-07 |
| GO_REGULATION_OF_INSULIN_SECRETION | 165 | 0.439 | 0.0422 | 0.092 | 9.11E-07 |
| GO_BETA_CATENIN_TCF_COMPLEX | 10 | 1.98 | 0.0471 | 0.424 | 1.48E-06 |
| GO_NEGATIVE_REGULATION_OF_TRANSCRIPTION_BY_RNA_POLYMERASE_II | 798 | 0.198 | 0.0412 | 0.0425 | 1.50E-06 |
| GO_DNA_BINDING_TRANSCRIPTION_FACTOR_ACTIVITY_RNA_POLYMERASE_II_SPECIFIC | 1011 | 0.183 | 0.0425 | 0.0394 | 1.69E-06 |
| GO_TRANSCRIPTION_FACTOR_BINDING | 613 | 0.222 | 0.0405 | 0.0484 | 2.35E-06 |
| GO_NEGATIVE_REGULATION_OF_BIOSYNTHETIC_PROCESS | 1435 | 0.146 | 0.0399 | 0.0322 | 2.82E-06 |
| GO_POSITIVE_REGULATION_OF_RNA_METABOLIC_PROCESS | 1579 | 0.14 | 0.04 | 0.031 | 2.96E-06 |
| GO_SIGNAL_RELEASE | 426 | 0.254 | 0.039 | 0.0571 | 4.31E-06 |
| GO_NEGATIVE_REGULATION_OF_RNA_BIOSYNTHETIC_PROCESS | 1160 | 0.157 | 0.0388 | 0.0358 | 5.90E-06 |
| GO_TRANSCRIPTION_REGULATOR_ACTIVITY | 1567 | 0.14 | 0.0397 | 0.032 | 6.45E-06 |
| GO_REGULATORY_REGION_NUCLEIC_ACID_BINDING | 892 | 0.175 | 0.0384 | 0.0407 | 8.37E-06 |
| GO_HEPATOCYTE_DIFFERENTIATION | 15 | 1.49 | 0.0433 | 0.346 | 8.52E-06 |
| GO_SEQUESTERING_OF_ZINC_ION | 8 | 2.27 | 0.0484 | 0.531 | 9.33E-06 |
| GO_DNA_BINDING_TRANSCRIPTION_FACTOR_ACTIVITY | 1228 | 0.153 | 0.0389 | 0.0364 | 1.28E-05 |
| GO_REGULATION_OF_PEPTIDE_HORMONE_SECRETION | 194 | 0.361 | 0.0376 | 0.0857 | 1.30E-05 |
| GO_LIPID_HOMEOSTASIS | 129 | 0.432 | 0.0367 | 0.103 | 1.34E-05 |
| GO_PEPTIDE_HORMONE_SECRETION | 234 | 0.323 | 0.0369 | 0.0772 | 1.45E-05 |
| GO_CIS_REGULATORY_REGION_BINDING | 573 | 0.214 | 0.0379 | 0.0513 | 1.52E-05 |
| GO_REGULATION_OF_HORMONE_SECRETION | 245 | 0.318 | 0.0371 | 0.0763 | 1.57E-05 |
| GO_HORMONE_TRANSPORT | 294 | 0.288 | 0.0368 | 0.0693 | 1.63E-05 |
| GO_HOMEOSTATIC_PROCESS | 1762 | 0.124 | 0.037 | 0.0301 | 2.03E-05 |
| GO_NEGATIVE_REGULATION_OF_NUCLEOBASE_CONTAINING_COMPOUND_METABOLIC_PROCESS | 1326 | 0.138 | 0.0363 | 0.0336 | 2.07E-05 |
| GO_DNA_BINDING_TRANSCRIPTION_FACTOR_BINDING | 334 | 0.258 | 0.0352 | 0.0641 | 2.77E-05 |
| GO_POSITIVE_REGULATION_OF_CELLULAR_BIOSYNTHETIC_PROCESS | 1811 | 0.117 | 0.0356 | 0.0292 | 2.92E-05 |
| GO_CHROMATIN_ORGANIZATION | 693 | 0.185 | 0.0359 | 0.0466 | 3.59E-05 |
| GO_DOUBLE_STRANDED_DNA_BINDING | 911 | 0.162 | 0.0359 | 0.0409 | 3.63E-05 |
| GO_CHROMOSOME_ORGANIZATION | 1082 | 0.146 | 0.0351 | 0.037 | 3.84E-05 |
| GO_DEVELOPMENTAL_GROWTH | 613 | 0.195 | 0.0356 | 0.0493 | 3.88E-05 |
| GO_REGULATION_OF_HORMONE_LEVELS | 491 | 0.213 | 0.035 | 0.055 | 5.35E-05 |
| GO_REPRESSING_TRANSCRIPTION_FACTOR_BINDING | 69 | 0.548 | 0.0342 | 0.142 | 5.74E-05 |
| GO_INNER_CELL_MASS_CELL_DIFFERENTIATION | 7 | 2.09 | 0.0417 | 0.543 | 5.78E-05 |
| GO_FAT_CELL_PROLIFERATION | 9 | 1.54 | 0.0348 | 0.406 | 7.28E-05 |
| GO_CHROMATIN | 1111 | 0.142 | 0.0345 | 0.0376 | 7.83E-05 |
| GO_POSITIVE_REGULATION_OF_TRANSCRIPTION_BY_RNA_POLYMERASE_II | 1108 | 0.138 | 0.0335 | 0.0368 | 8.45E-05 |
| GO_OLFACTORY_BEHAVIOR | 9 | 1.34 | 0.0302 | 0.361 | 0.000102 |
| GO_REGULATION_OF_EPITHELIAL_CELL_MIGRATION | 208 | 0.315 | 0.034 | 0.0848 | 0.000102 |
| GO_CELL_FATE_COMMITMENT | 252 | 0.291 | 0.0345 | 0.0784 | 0.000104 |
| GO_REGULATION_OF_DEVELOPMENTAL_GROWTH | 306 | 0.255 | 0.0333 | 0.0693 | 0.000115 |
| GO_CELLULAR_RESPONSE_TO_CARBOHYDRATE_STIMULUS | 130 | 0.371 | 0.0317 | 0.101 | 0.00012 |
| GO_VENTRICULAR_SEPTUM_MORPHOGENESIS | 40 | 0.776 | 0.0369 | 0.212 | 0.000123 |
| GO_ADENYLATE_CYCLASE_ACTIVATING_DOPAMINE_RECEPTOR_SIGNALING_PATHWAY | 10 | 1.57 | 0.0373 | 0.431 | 0.000133 |
| GO_BLOOD_VESSEL_ENDOTHELIAL_CELL_MIGRATION | 106 | 0.416 | 0.0321 | 0.114 | 0.000139 |
| GO_CELLULAR_GLUCOSE_HOMEOSTASIS | 136 | 0.359 | 0.0314 | 0.0987 | 0.000139 |
| GO_IN_UTERO_EMBRYONIC_DEVELOPMENT | 355 | 0.229 | 0.0321 | 0.063 | 0.000142 |
| GO_NUCLEAR_CHROMOSOME | 1152 | 0.134 | 0.033 | 0.0371 | 0.000156 |
| GO_SEQUENCE_SPECIFIC_DNA_BINDING | 1098 | 0.136 | 0.0329 | 0.0378 | 0.000158 |
| GO_PROTEIN_SERINE_THREONINE_KINASE_INHIBITOR_ACTIVITY | 29 | 0.855 | 0.0346 | 0.239 | 0.000174 |
| GO_OXIDATIVE_RNA_DEMETHYLATION | 5 | 2.34 | 0.0393 | 0.657 | 0.000186 |
| GO_OXIDATIVE_RNA_DEMETHYLASE_ACTIVITY | 5 | 2.34 | 0.0393 | 0.657 | 0.000186 |
| GO_CHEMOSENSORY_BEHAVIOR | 15 | 1.03 | 0.03 | 0.29 | 0.000191 |
| GO_SEQUENCE_SPECIFIC_DOUBLE_STRANDED_DNA_BINDING | 824 | 0.152 | 0.032 | 0.0429 | 0.000202 |
| GO_CIRCADIAN_SLEEP_WAKE_CYCLE_NON_REM_SLEEP | 9 | 1.5 | 0.0339 | 0.425 | 0.000203 |
| GO_CHEMICAL_HOMEOSTASIS | 1089 | 0.132 | 0.0316 | 0.0378 | 0.000245 |
| GO_NEGATIVE_REGULATION_OF_GROWTH | 224 | 0.28 | 0.0313 | 0.0803 | 0.000248 |
| GO_DNA_APURINIC_OR_APYRIMIDINIC_SITE_ENDONUCLEASE_ACTIVITY | 11 | 1.18 | 0.0294 | 0.339 | 0.000256 |
| GO_TISSUE_MIGRATION | 274 | 0.257 | 0.0317 | 0.0739 | 0.00026 |
| GO_EMBRYO_DEVELOPMENT_ENDING_IN_BIRTH_OR_EGG_HATCHING | 613 | 0.171 | 0.0314 | 0.0496 | 0.000273 |
| GO_INSULIN_BINDING | 5 | 1.96 | 0.0329 | 0.568 | 0.00029 |
| GO_NEGATIVE_REGULATION_OF_SOMATIC_STEM_CELL_POPULATION_MAINTENANCE | 4 | 2.39 | 0.0359 | 0.703 | 0.000338 |
| GO_AT_DNA_BINDING | 9 | 1.32 | 0.0298 | 0.394 | 0.000391 |
| GO_POSITIVE_REGULATION_OF_INSULIN_SECRETION | 68 | 0.452 | 0.028 | 0.136 | 0.000426 |
| GO_NEGATIVE_REGULATION_OF_TRIGLYCERIDE_BIOSYNTHETIC_PROCESS | 2 | 3.33 | 0.0355 | 1 | 0.000428 |
| GO_PURINERGIC_RECEPTOR_SIGNALING_PATHWAY | 28 | 0.864 | 0.0343 | 0.261 | 0.000469 |
| GO_POTASSIUM_ION_IMPORT_ACROSS_PLASMA_MEMBRANE | 46 | 0.646 | 0.0329 | 0.196 | 0.000495 |
| GO_CELL_PROLIFERATION_IN_BONE_MARROW | 8 | 1.24 | 0.0265 | 0.38 | 0.000533 |
| GO_EMBRYO_DEVELOPMENT | 966 | 0.131 | 0.0298 | 0.0401 | 0.000546 |
| GO_REGULATION_OF_RESPIRATORY_SYSTEM_PROCESS | 14 | 1.05 | 0.0296 | 0.323 | 0.000557 |
| GO_LBD_DOMAIN_BINDING | 6 | 1.55 | 0.0286 | 0.479 | 0.000588 |
| GO_CHROMATIN_ORGANIZATION_INVOLVED_IN_REGULATION_OF_TRANSCRIPTION | 106 | 0.412 | 0.0318 | 0.127 | 0.000591 |
| GO_RESPONSE_TO_ZINC_ION | 48 | 0.612 | 0.0319 | 0.19 | 0.000641 |
| GO_VERY_LOW_DENSITY_LIPOPROTEIN_PARTICLE_ASSEMBLY | 11 | 1.23 | 0.0307 | 0.383 | 0.000656 |
| GO_INTESTINAL_ABSORPTION | 37 | 0.596 | 0.0272 | 0.186 | 0.000672 |
| GO_ACYLGLYCEROL_ACYL_CHAIN_REMODELING | 5 | 1.56 | 0.0262 | 0.486 | 0.000689 |
| GO_REGULATION_OF_HEART_GROWTH | 63 | 0.527 | 0.0314 | 0.165 | 0.000693 |
| GO_SKELETAL_MUSCLE_THIN_FILAMENT_ASSEMBLY | 9 | 1.4 | 0.0316 | 0.439 | 0.000716 |
| GO_RESPONSE_TO_LEPTIN | 22 | 1.01 | 0.0356 | 0.318 | 0.000748 |
| GO_MEMBRANE_REPOLARIZATION_DURING_ATRIAL_CARDIAC_MUSCLE_CELL_ACTION_POTENTIAL | 5 | 1.92 | 0.0322 | 0.603 | 0.000751 |
| GO_TRANSCRIPTION_INITIATION_FROM_RNA_POLYMERASE_II_PROMOTER | 170 | 0.298 | 0.029 | 0.0941 | 0.00078 |
| GO_CARDIAC_SEPTUM_MORPHOGENESIS | 72 | 0.495 | 0.0315 | 0.156 | 0.00078 |
| GO_NEUTRAL_LIPID_BIOSYNTHETIC_PROCESS | 34 | 0.614 | 0.0269 | 0.194 | 0.000783 |
| GO_ENDOCARDIUM_MORPHOGENESIS | 6 | 1.93 | 0.0355 | 0.612 | 0.00082 |
| GO_KINASE_BINDING | 684 | 0.143 | 0.0277 | 0.046 | 0.00092 |
| GO_GROWTH | 902 | 0.126 | 0.0278 | 0.0405 | 0.000926 |
| GO_REGULATION_OF_CELL_CYCLE | 1075 | 0.114 | 0.0272 | 0.0368 | 0.000982 |
| GO_PEPTIDE_HORMONE_BINDING | 49 | 0.573 | 0.0301 | 0.185 | 0.001011 |
| GO_FIBROBLAST_GROWTH_FACTOR_BINDING | 23 | 0.784 | 0.0283 | 0.255 | 0.001051 |
| GO_NEGATIVE_REGULATION_OF_CHROMATIN_SILENCING | 17 | 1.07 | 0.0331 | 0.347 | 0.001052 |
| GO_OTIC_VESICLE_MORPHOGENESIS | 10 | 1.57 | 0.0373 | 0.511 | 0.001063 |
| GO_PULMONARY_VALVE_DEVELOPMENT | 21 | 0.823 | 0.0283 | 0.268 | 0.001074 |
| GO_MOTOR_NEURON_MIGRATION | 6 | 1.76 | 0.0325 | 0.575 | 0.001074 |
| GO_NEGATIVE_REGULATION_OF_RNA_POLYMERASE_II_REGULATORY_REGION_SEQUENCE_SPECIFIC_DNA_BINDING | 5 | 1.99 | 0.0334 | 0.648 | 0.001075 |
| GO_VASCULAR_ASSOCIATED_SMOOTH_MUSCLE_CELL_APOPTOTIC_PROCESS | 9 | 1.18 | 0.0267 | 0.386 | 0.001101 |
